# The glymphatic system clears amyloid beta and tau from brain to plasma in humans

**DOI:** 10.1101/2024.07.30.24311248

**Authors:** Paul Dagum, Donald L. Elbert, Laurent Giovangrandi, Tarandeep Singh, Venky V. Venkatesh, Alejandro Corbellini, Robert M. Kaplan, Swati Rane Levendovszky, Elizabeth Ludington, Kevin Yarasheski, Jeffrey Lowenkron, Carla VandeWeerd, Miranda M. Lim, Jeffrey J. Iliff

## Abstract

Poor sleep is implicated in the development of Alzheimer’s disease (AD) pathology and cognitive impairment. Dysfunction of the glymphatic system has been proposed as a mechanistic link between sleep disruption and AD, and in animal models glymphatic impairment is sufficient to drive the development of AD pathology. It remains unknown whether the glymphatic system clears amyloid beta (Aβ) and tau from the brain in humans. This study directly tested the hypothesis that sleep-active glymphatic clearance increases morning plasma AD biomarker levels. In a multi-site randomized crossover clinical trial, participants (N=39) underwent overnight in-laboratory conditions of normal sleep and sleep deprivation following instrumentation that included a novel device to measure sleep features by electroencephalography (EEG), cerebrovascular compliance by measurement of pulse transit time (PTT), and brain parenchymal resistance to glymphatic flow (R_P_) by transcranial multifrequency impedance spectroscopy. We evaluated whether R_P_, sleep EEG features, PTT, and heart rate variability (HRV) predicted overnight changes in plasma AD biomarker (Aβ40, Aβ42, np-tau181, np-tau217 and p-tau181) levels. We found that changes in R_P_, PTT, HRV and EEG delta power in a multivariate linear mixed model predicted sleep-associated overnight changes in Aβ40 (p=0.013), Aβ42 (p<0.001), np-tau181 (p<0.001), np-tau217 (p<0.001) and p-tau181 (p<0.001). The predicted changes replicated those from a multicompartment model based on published data on synaptic-metabolic release and clearance of Aβ and tau to the plasma. Sensitivity analysis demonstrated that while morning biomarker levels were altered by both the synaptic-metabolic release and clearance of Aβ and tau, during sleep glymphatic clearance was the dominant influence. In contrast, during sleep deprivation (waking) while glymphatic clearance was a contributor, Aβ and tau release was the dominant factor explaining variance in morning plasma Aβ and tau levels. Our findings show that elements of sleep-active physiology, in particular decreased brain parenchymal resistance, facilitates the overnight clearance of AD biomarkers to plasma, supporting a role for glymphatic clearance in these processes, and suggesting the enhancement of glymphatic function as a therapeutic target to reduce the development and progression of AD pathology in at-risk populations.

## INTRODUCTION

Sleep disruption is associated with increased risk of Alzheimer’s disease (AD) in epidemiological studies^1–3^, while biomarker studies in cognitively-intact participants suggest that short sleep duration and poor sleep quality are associated with greater AD-related amyloid β (Aβ) and tau pathology^4–9^, even before the onset of clinical symptoms of cognitive and functional decline. Yet the mechanistic link between sleep disruption and AD risk remains undefined. The glymphatic system is a brain-wide network of perivascular pathways along which cerebrospinal fluid (CSF) and interstitial fluid (ISF) exchange, supporting the clearance of interstitial solutes from the brain into the CSF, and thence into the blood. Studies in mice demonstrated that glymphatic function contributes to the clearance of Aβ and tau, is more rapid during sleep, and is impaired by acute sleep deprivation^10–15^. While more rapid glymphatic clearance of solutes during sleep has now been validated in the human brain^16,17^, no study has tested mechanistically whether sleep-active glymphatic activity contributes to the clearance of Aβ and tau in a healthy human population.

Plasma biomarkers have emerged as a potentially critical tool enabling the non-invasive diagnosis of AD at a large scale within the wider population. Immuno-assay and mass spectrometry-based measures of different Aβ and tau species permit the assessment of AD-related Aβ plaque burden and tau pathology, even during preclinical stages of disease in the years prior to patients’ clinical progression to cognitive impairment and dementia^18,19^. Although generally regarded as steady-state indicators of brain AD pathological burden, plasma AD biomarker levels are impacted acutely by sleep disruption^4,20^. While it is possible that these effects reflect reduced rates of cellular Aβ and tau release during sleep, they may also reflect the role of sleep-active glymphatic clearance in the transport of Aβ and tau from the brain to the blood. In a crossover study including 5 healthy participants, plasma Aβ_40_, Aβ_42_, non-phosphorylated tau181 (np-tau181), non-phosphorylated tau217 (np-tau217) and phosphorylated tau181 (p-tau181) were each reduced during the overnight period by acute sleep deprivation^4^. Noting an increased ratio of CSF:plasma Aβ and tau levels, the authors inferred that the processes governing the clearance of Aβ and tau from the brain to the CSF, and thence from the CSF to the plasma must be impaired by acute sleep deprivation. These findings are consistent with the sleep-active glymphatic clearance of Aβ and tau from the brain, yet they do not provide causal support. A second study showed that plasma Aβ and tau levels were reduced in participants that exhibited MRI evidence of impaired glymphatic and CSF-to-plasma clearance^21^. These findings established a correlation between steady-state AD biomarker plasma levels and impaired CSF circulation, but did not test whether sleep-active glymphatic function directly contributes to the clearance of these peptides during sleep in otherwise healthy individuals.

Here we designed a study to directly test if sleep-active glymphatic function contributes to the overnight clearance of Aβ and tau from the human brain. We first developed a compartmental pharmacokinetic model to predict the effect that sleep-associated changes in Aβ and tau release and sleep-active glymphatic clearance, reflected by transport between the brain ISF and CSF, would have on morning plasma levels of Aβ and tau species. We then carried out a clinical randomized crossover study where participants underwent overnight normal sleep and sleep deprivation. Prior studies in rodents and humans demonstrate that glymphatic function increases with increasing sleep electroencephalography (EEG) delta and theta power, reduced EEG beta power, reduced heart rate (HR), reduced central noradrenergic tone, and reduced brain parenchymal resistance to fluid flow (R_P_), and vasomotor oscillations^13,22–26^. Measuring these key determinants of glymphatic exchange with a novel investigational device^17^, we defined the relationship between sleep features measured by EEG, R_P_ measured by dynamic impedance spectroscopy, and HRV and cerebrovascular compliance measured by impedance plethysmography (IPG) and overnight changes in plasma AD biomarker levels. Using this approach, we directly tested the hypothesis that sleep-active glymphatic clearance elevates morning plasma AD biomarker levels, despite a sleep-associated reduction in their production.

## RESULTS

Statistical significance tests presented in the results have not been adjusted for multiple comparisons.

### Compartmental pharmacokinetic model relating glymphatic exchange to plasma AD biomarker levels

We first developed a compartmental pharmacokinetic model to define the effects that sleep-related changes in interstitial solute release and clearance would have on overnight changes in plasma levels of Aβ and tau. The model (shown in **Figure 1A**) is a simplification of prior models^27^ and includes the minimal number of compartments, transport and elimination pathways needed to define the effect that a change in overnight interstitial solute production/release and glymphatic clearance between the ISF and CSF would have on plasma Aβ and tau levels. The model included a 16-hour period of wake, and either an 8-hour period of sleep or an 8-hour period of sleep deprivation. It includes 6 compartments: Cellular Site of Production/Release (**1**), Free ISF (**2**), CSF (**3**), Plasma (**4**), Cellular Site of Uptake/Degradation (**5**), and Non-Monomeric Pools (**6**). Solute transport processes and their rate constants for Aβ and tau species based on literature values^27,28^ are provided in **Table 1**.

**Table 1:**
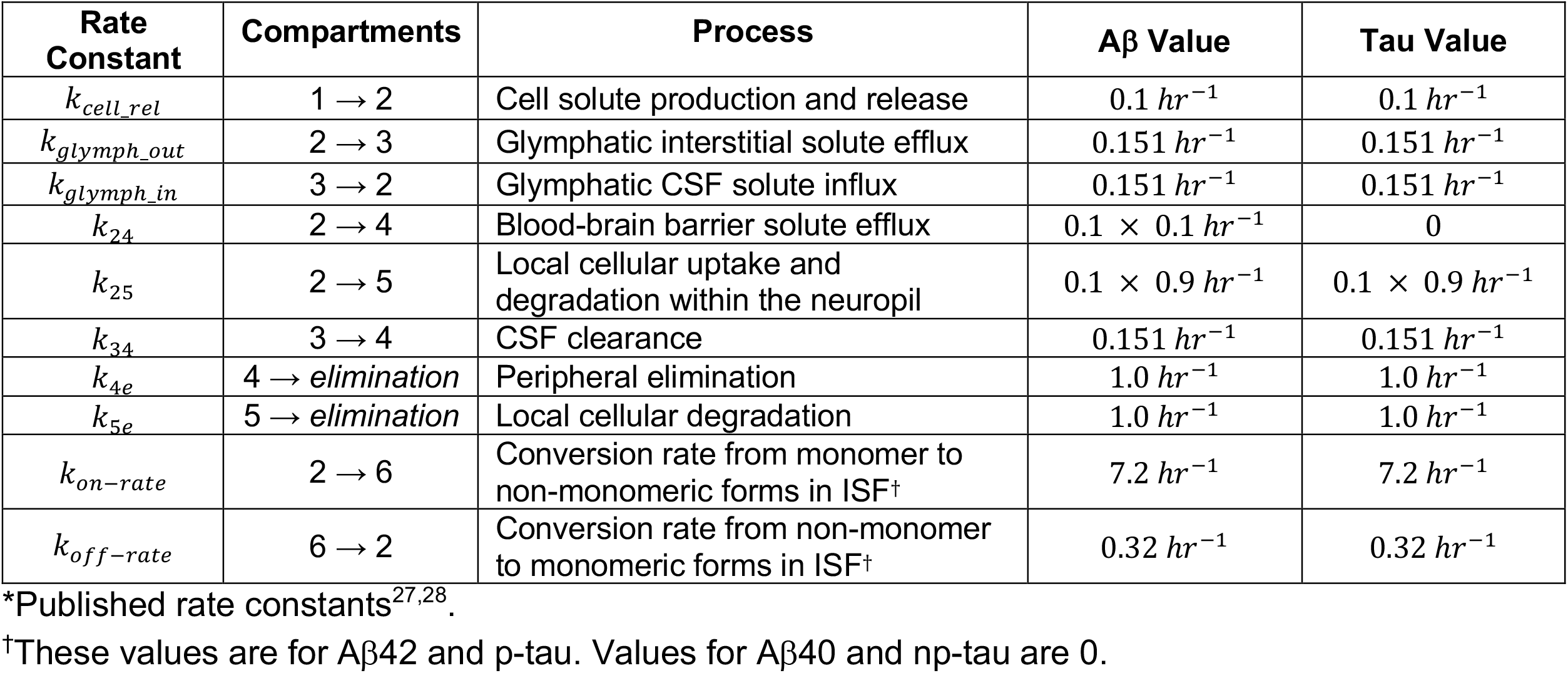
Compartment rate constants of amyloid β and tau transport*.

| Rate Constant | Compartments | Process | Aβ Value | Tau Value |
| --- | --- | --- | --- | --- |
| $k_{cell\_rel}$ | $1 \rightarrow 2$ | Cell solute production and release | $0.1 \text{ hr}^{-1}$ | $0.1 \text{ hr}^{-1}$ |
| $k_{glymph\_out}$ | $2 \rightarrow 3$ | Glymphatic interstitial solute efflux | $0.151 \text{ hr}^{-1}$ | $0.151 \text{ hr}^{-1}$ |
| $k_{glymph\_in}$ | $3 \rightarrow 2$ | Glymphatic CSF solute influx | $0.151 \text{ hr}^{-1}$ | $0.151 \text{ hr}^{-1}$ |
| $k_{24}$ | $2 \rightarrow 4$ | Blood-brain barrier solute efflux | $0.1 \times 0.1 \text{ hr}^{-1}$ | 0 |
| $k_{25}$ | $2 \rightarrow 5$ | Local cellular uptake and degradation within the neuropil | $0.1 \times 0.9 \text{ hr}^{-1}$ | $0.1 \times 0.9 \text{ hr}^{-1}$ |
| $k_{34}$ | $3 \rightarrow 4$ | CSF clearance | $0.151 \text{ hr}^{-1}$ | $0.151 \text{ hr}^{-1}$ |
| $k_{4e}$ | $4 \rightarrow \text{elimination}$ | Peripheral elimination | $1.0 \text{ hr}^{-1}$ | $1.0 \text{ hr}^{-1}$ |
| $k_{5e}$ | $5 \rightarrow \text{elimination}$ | Local cellular degradation | $1.0 \text{ hr}^{-1}$ | $1.0 \text{ hr}^{-1}$ |
| $k_{on-rate}$ | $2 \rightarrow 6$ | Conversion rate from monomer to non-monomeric forms in ISF<sup>†</sup> | $7.2 \text{ hr}^{-1}$ | $7.2 \text{ hr}^{-1}$ |
| $k_{off-rate}$ | $6 \rightarrow 2$ | Conversion rate from non-monomer to monomeric forms in ISF<sup>†</sup> | $0.32 \text{ hr}^{-1}$ | $0.32 \text{ hr}^{-1}$ |

\*Published rate constants<sup>27,28</sup>.
<sup>†</sup>These values are for Aβ42 and p-tau. Values for Aβ40 and np-tau are 0.

**Figure 1.**
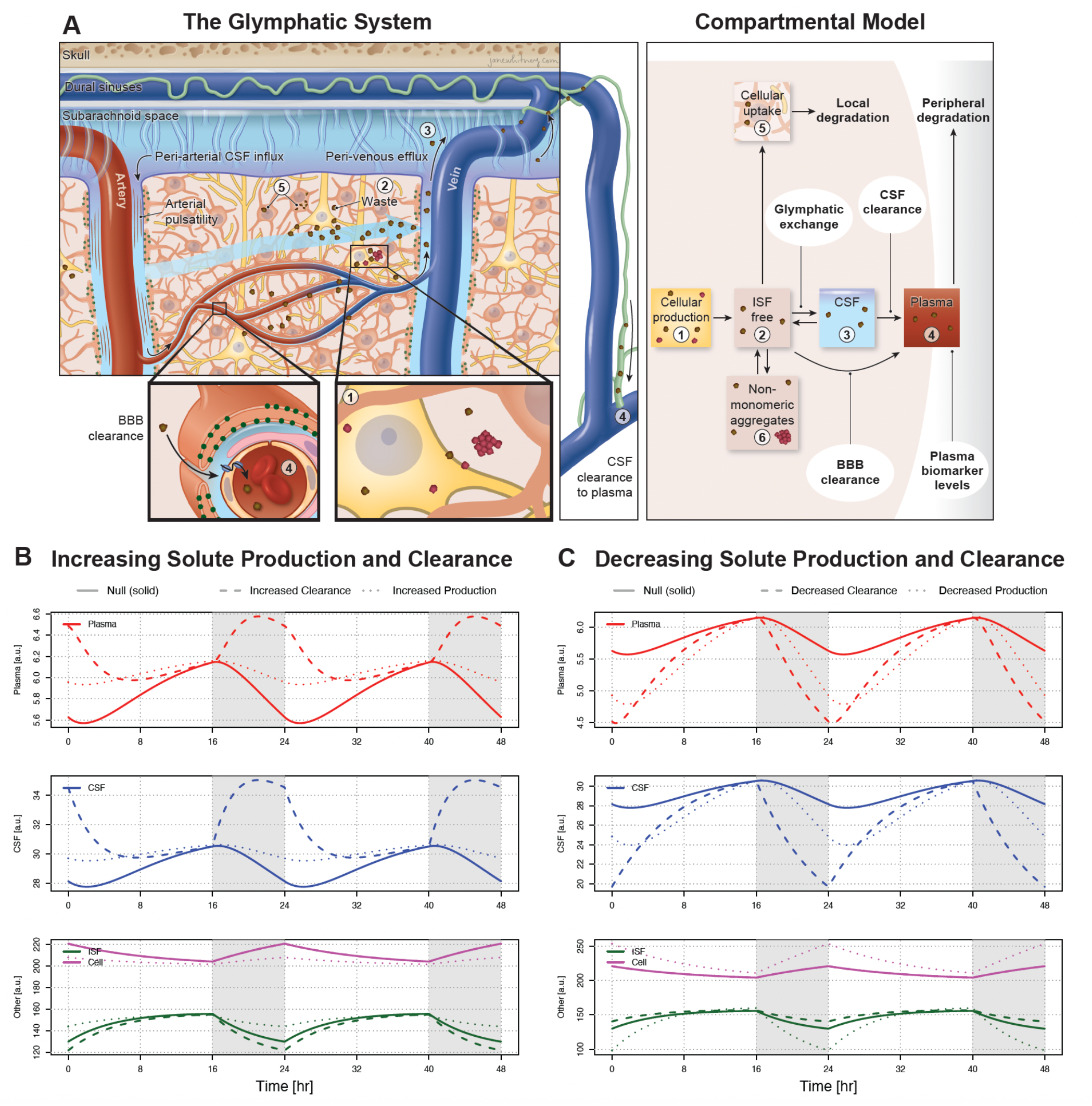
Six-compartment model of brain to plasma solute exchange. **(A)** Amyloid β (Aβ) and tau are produced in neurons (**1**) and released into the ISF compartment (**2**) where they are cleared from the ISF via local cellular uptake (**2** ➜ **5**) and degradation, blood-brain barrier efflux (**2** ➜ **4**), or glymphatic efflux to the CSF (**2** ➜ **3**). Aβ42 and phosphorylated tau species are prone to non-monomeric aggregation, unlike Aβ40 and non-phosphorylated tau. Their monomeric forms can also be cleared from the ISF through further aggregation into non-monomeric structures (**2** ➜ **6**). CSF solutes may recirculate back into the brain interstitium (**3** ➜ **2**) or be cleared by CSF efflux pathways to the plasma (**3** ➜ **4**) from whence peripheral degradation occurs. **(B) – (C)** Compartment concentrations of monomeric amyloid β and tau at steady state in the **null model** and the **neuro-glymphatic model** following changes in glymphatic efflux/influx or synaptic and metabolic release. Release of cellular Aβ and tau species from neurons (**1**) into the ISF (**2**) in the model is controlled by the cellular solute release rate constant *k*_*cell*_*rel*_ and was kept constant during the 16 hours of wake and was reduced by 30% during the 8 hours of sleep occurring in the shaded window between16 and 24 hours. Solid lines show compartment concentrations under the **null model**, with time-invariant release rate constant *k*_*cell*_*rel*_ and time-invariant exchange between the CSF (**3**) and ISF (**2**) compartments. Long dashed lines show change in compartment concentrations when glymphatic efflux/influx rate constants *k*_*glymph*_*out*_ and *k*_*glymph*_*in*_, were increased by a factor of 1.5 **(B)** or decreased by a factor 0.5 **(C)** during the 8-hour sleep window. Dotted lines show change in compartment concentrations when synaptic and metabolic release rate constant *k*_*cell*_*rel*_ was increased by a factor of 1.25 **(B)** or decreased by a factor of 0.5 **(C)** during the 8-hour sleep window. Note that increasing (decreasing) either glymphatic efflux/influx or synaptic and metabolic release during sleep increases (decreases) morning plasma level of Aβ and tau species relative to the **null model**.

The 24-hour cellular synthesis of amyloid precursor protein (APP) and tau is invariant to sleep-wake state and was modeled at a constant rate of 10 arbitrary units (a.u.) per hour^29,30^. Release of cellular Aβ and tau species into the ISF in the model is controlled by the cellular solute release rate constant *k*_*cell*_*rel*_ (**Table 1**) and was kept constant during the 16 hours of wake. During the 8 hours of sleep, *k*_*cell*_*rel*_ was reduced by 30% consistent with reported 30% sleep reduction in ISF Aβ and tau species^29,30^, and during the 8 hours of sleep deprivation it was reduced by 10% reflecting a circadian time-of-day effect.

This model was evaluated under two conditions. In the **null model**, sleep-active glymphatic exchange was posited to not occur; therefore solute transport between the Free ISF compartment (**2**) and the CSF compartment (**3**), reflected by the rate constants *k*_*glymph*_*out*_ and *k*_*glymph*_*in*_, were held constant across both sleep and waking (or sleep deprivation) states. In this model, sleep-related changes in synaptic and metabolic activity, which impact cellular solute release, were not permitted to vary beyond the defined reduction in the cellular solute release rate constant *k*_*cell*_*rel*_ during the 8-hour sleep period. In the **neuro-glymphatic model**, *k*_*glymph*_*out*_ and *k*_*glymph*_*in*_ increased during sleep, reflecting the sleep-active CSF solute influx and interstitial solute efflux observed in both rodents and humans^13,16,17^. Additionally, *k*_*cell*_*rel*_ either increased or decreased with sleep, representing corresponding changes in synaptic and metabolic activity that affect solute release into the brain interstitium^30,31^.

The full derivation of the compartmental pharmacokinetic model is provided in **Supplementary Methods and Results. Figure 1B** presents the steady-state compartmental concentrations of Aβ and tau over a 48-hour period under the **null model** (solid lines), as well as under **neuro-glymphatic model** scenarios involving increased glymphatic clearance (dashed lines) and elevated synaptic-metabolic release (dotted lines). **Figure 1C** illustrates alternative **neuro-glymphatic model** scenarios with reduced glymphatic clearance (dashed lines) and decreased synaptic-metabolic release (dotted lines), shown in comparison to the **null model** (solid lines). Across all scenarios, both Aβ and tau concentrations in the plasma increase or decrease relative to the **null model**, depending on whether glymphatic clearance or synaptic-metabolic release is enhanced or diminished, respectively, over the 48-hour period. These plots demonstrate that changes in Aβ and tau concentrations within the plasma and CSF compartments cannot independently distinguish between decreased glymphatic clearance and decreased solute release, nor between increased glymphatic clearance and increased solute release. An additional derivation of the model, described in detail in the **Supplementary Methods and Results**, demonstrates that within the in the **null model**, morning plasma levels will be linearly dependent on evening plasma levels only, whereas the **neuro-glymphatic model** has a contribution from the interaction term between the evening plasma levels and the variable rate constants *k*_*glymph*_*out*_, *k*_*glymph*_*in*_ and *k*_*cell*_*rel*_.

We next compared changes in plasma levels of species that remained primarily monomeric in form (Aβ40, np-tau), and species that are prone to form non-monomeric aggregates (Aβ42, p-tau) following increases and decreases in glymphatic clearance and synaptic-metabolic release. If this tendency to form aggregates is ignored, and rate constants for monomer-to-aggregate conversion is set to zero (*k*_*on*-*rate*_= 0), changes in cellular release *k*_*cell*_*rel*_ or clearance *k*_*glymph*_*out*_ have no effect on the Aβ42/Aβ40 or p-tau/np-tau ratios (**Supplemental Table 1**, top row). In contrast, when the tendency for Aβ42 and p-tau to form non-monomeric aggregates is included in the model (*k*_*on*-*rate*_> 0), increased and decreased solute release and glymphatic clearance shift plasma Aβ42/Aβ40 or p-tau/np-tau ratios (**Supplemental Table 1**). Both the Aβ42/Aβ40 or p-tau/np-tau ratios increase in response to decreased production or increased glymphatic clearance, and decrease in response to increased production or reduced clearance. This suggests that non-monomeric aggregates act both as a secondary source of monomeric Aβ42 and p-tau species under conditions of reduced production and as an additional sink for these species under conditions of reduced clearance. Under conditions of increased glymphatic clearance, the enhanced removal of Aβ42 and p-tau from the ISF reduces their opportunity to aggregate, resulting in a higher proportion reaching the plasma relative to Aβ40 and np-tau—thus increasing their respective plasma ratios. Taken together, this suggests that changes in plasma Aβ42/Aβ40 or p-tau/np-tau ratios allow us to infer the predominant mechanism driving changes in morning plasma AD biomarker levels under sleep or sleep-deprivation conditions in experimental settings.

These pharmacokinetic modeling results provide three concrete predictions that permit us to test the hypothesis that sleep-active glymphatic exchange contributes to overnight changes in plasma AD biomarker levels independent of sleep-dependent changes in solute release:

1. Under the **neuro-glymphatic model**, compared to normal sleep, decreased clearance during overnight sleep deprivation would reduce morning plasma AD biomarker levels while increased production would increase levels.
2. Under the **neuro-glymphatic model**, under conditions of sleep deprivation, morning plasma AD biomarker levels would increase by the interaction term between evening plasma levels and features of synaptic-metabolic activity contributing to greater production; under normal sleep conditions morning plasma AD biomarker levels will be increased by the interaction term between evening plasma levels and features of sleep-active glymphatic function contributing to greater clearance.
3. Under the **neuro-glymphatic model**, under normal sleep conditions morning plasma Aβ42/Aβ40 and p-tau/np-tau ratios will increase with greater sleep-dependent clearance; overnight sleep deprivation will decrease these ratios from greater wake-dependent production.

### Clinical study design and participant demographic data

As described previously^17^, we conducted two cross-over clinical studies in which participants underwent one night of normal sleep and one night of sleep deprivation, in randomized order and separated by two or more weeks (**Figure 2A**). One study was conducted in The Villages® community in Central Florida where the University of Florida maintains a satellite academic research center, The UF Health Precision Health Research Center (UF Health PHRC). The second study was carried out at the University of Washington (UW) in Seattle. Study participants underwent peripheral blood draws at 1900 hrs and 0700 hrs, prior to and following the overnight sleep and sleep deprivation periods. Plasma AD biomarkers, including Aβ_40_, Aβ_42_, np-tau181, np-tau217, p-tau181, and phosphorylated tau217 (p-tau217) were quantified using C_2_N Diagnostics’ immunoprecipitation liquid chromatography-tandem mass spectrometry platforms^32–34^. Details of specimen collection and processing are provided in **Supplementary Methods and Results**. During the overnight period, participants were instrumented with an investigational in-ear wearable device that measured key determinants of glymphatic function, including sleep features (hypnogram and spectral band power) by electroencephalography (EEG), heart rate variability (HRV) by photoplethysmography (PPG), cerebrovascular pulse transit time (PTT) by impedance plethysmography (IPG), and brain parenchymal resistance (R_P_) by dynamic impedance spectroscopy (**Figure 2B**). A detailed description of this device, the validation of its sleep EEG measures against gold-standard overnight polysomnography, and of the measures of R_P_ against contrast-enhanced magnetic resonance imaging (CE-MRI)-based measures of glymphatic function has recently been reported^17^. Cerebrovascular PTT measurements obtained using impedance plethysmography were validated against contrast-enhanced MRI-based measures of cerebrovascular function, as detailed in the **Supplemental Methods and Results (Supplemental Tables 2 – 6)**.

**Figure 2.**
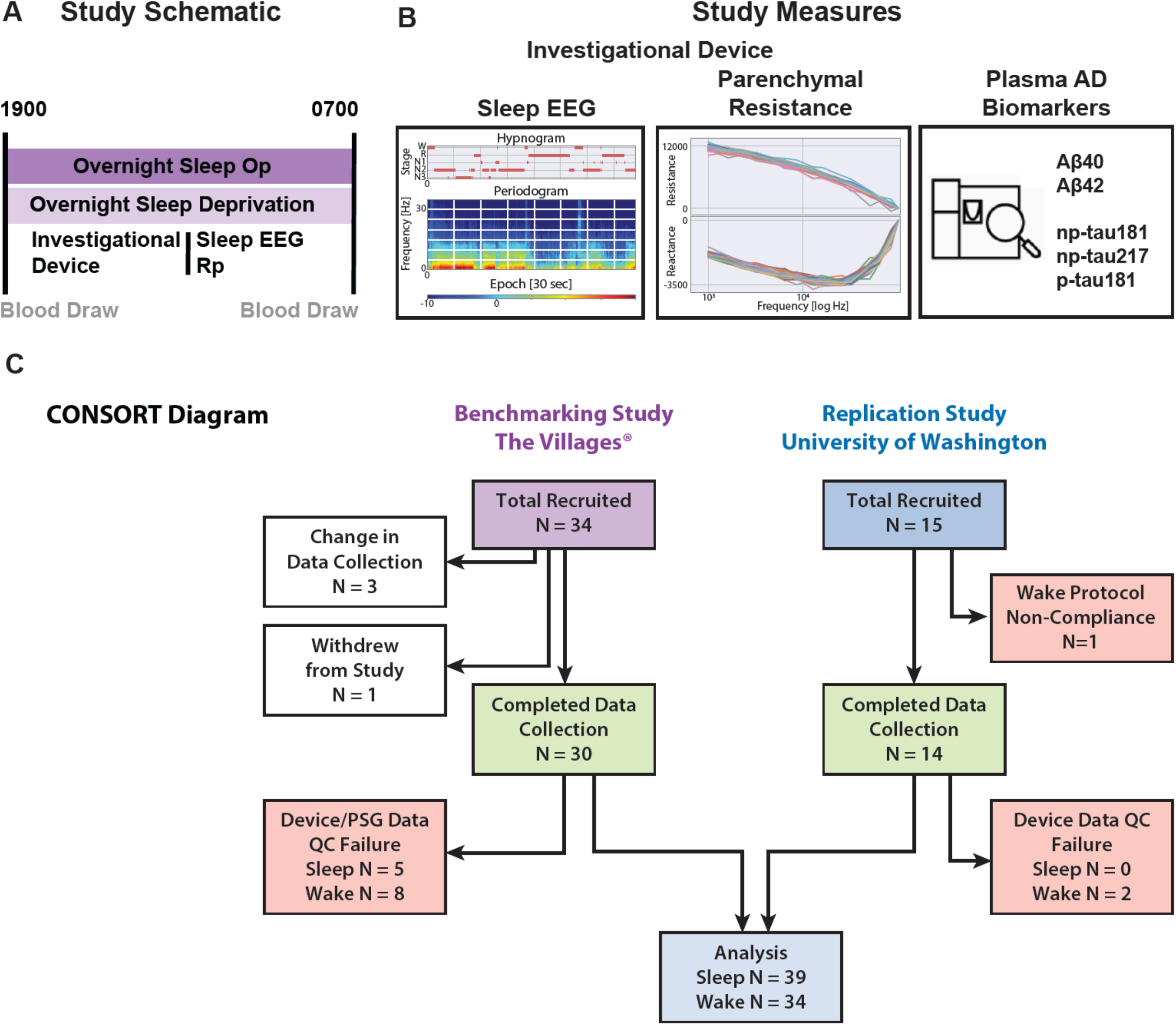
Study schematic and CONSORT diagram. The Benchmarking Study conducted at The Villages® and the Replication study conducted at the University of Washington were **(A)** randomized cross-over assignment of overnight sleep opportunity and overnight sleep deprivation designed to define the relationship between parenchymal resistance (R_P_) and glymphatic function. **(B)** Reported here are the overnight investigational device recordings of R_P_, EEG and HR, and blood analysis of amyloid β and tau levels (Aβ40, Aβ42, np-tau181, np-tau217 and p-tau181). **(C)** The Benchmarking Study enrolled 34 participants of which 30 completed both visits. Three were censored due to changes in device data collection and sensor locations. One withdrew following the first MRI scan. Of the 30 that completed the study, 5 overnight sleep visits and 8 overnight wake visits failed the data quality control (QC) criteria to provide sufficient artifact free data to yield results. This resulted in 25 sleep and 22 wake complete data sets. The Replication Study enrolled 14 participants. All 14 completed the study, of which 3 wake visits failed the data QC criteria and one sleep visit failed specific to the analysis of these data because of missing EEG powerband data during REM sleep caused by artifacts. The remaining 13 participants had complete data for analysis.

All studies were performed between October 2022 - June 2023 and were reviewed and approved by University of Florida Institutional Review Board (IRB No. 202201364, Villages study) and Western Institutional Review Board (IRB No. 20225818, UW study). The studies have been registered at ClinicalTrials.gov (https://clinicaltrials.gov/study/NCT06060054 and https://clinicaltrials.gov/study/NCT06222385). Written informed consent was obtained from all study participants during a screening visit, prior to any study activities. Studies were carried out in accordance with the principles of the Belmont Report. The Villages Study enrolled 34 healthy participants 56-66 years of age. The UW Study enrolled 14 healthy participants 49-63 years of age. Participants were excluded if they had cognitive impairment or clinical depression. Cognitive impairment was assessed using the Montreal Cognitive Assessment (MoCA, 28.1 +/− 1.2; range 26, 30) and depression was evaluated using the 15-item Geriatric Depression Scale (GDS, 0.7 +/− 1.2; range 0, 4). Participants with a self-reported history of diabetes, hypertension, coronary artery disease, pulmonary disease, neurological disease, depression, or anxiety were excluded from the study. Exclusion also applied to participants planning travel to alternate time zones within two weeks of study participation, as well as those with a formal diagnosis of any sleep disorder (e.g., sleep apnea requiring positive airway pressure [PAP] therapy, insomnia, restless leg syndrome, circadian rhythm sleep disorder, or parasomnia). To minimize preparation bias (i.e., altering sleep schedules or taking naps in anticipation of sleep deprivation visits), participants were not informed of their initial visit assignment until 4:00 PM on the day of arrival. Sleep quality prior to and between study visits was subjectively evaluated based on investigator assessment. Participant demographics, MoCA and GDS scores are listed for each study site and for the combined dataset in **Table 2**.

**Table 2.** Clinical study demographic information*.

| **Study** | **Age** | **Sex**<br>**(F/M)** | **APOE-e4**<br>**(+/-)** | **Amyloid**<br>**Plaque**<br>**Status**<br>**(+/-)** | **MoCA** | **GDS** |
| --- | --- | --- | --- | --- | --- | --- |
| **Villages** | 61.9 (2.7) | 13/12 | 7/18 | 11/14 | 27.9 (1.2) | 0.7 (1.2) |
| **University of**<br>**Washington** | 55.9 (4.6) | 6/7 | 4/9 | 0/13 | 28.5 (1.2) | 0.5 (1.1) |
| **Combined** | 59.8 (4.5) | 19/19 | 11/27 | 11/27 | 28.1 (1.2) | 0.7 (1.2) |

\*Values are Mean (SD) or count. MoCA, Montreal Cognitive Assessment; GDS, Geriatric Depression Scale. Amyloid plaque status determined using C<sub>2</sub>N PrecivityAD test with cutoff of 0.089<sup>32,34,35</sup>.

A Consolidated Standards of Reporting Trials (CONSORT) diagram for the Villages Study and UW Study is provided in **Figure 2C**. Within the Villages Study, the first three participants were removed from analysis because of a sensor position change in the investigational device. One participant was unable to complete the first MRI session and withdrew from the study. Of the remaining 30 participants (61.8 ± 2.7 years of age; 14 female, 16 male) that completed the Villages Study, five overnight sleep studies and eight overnight wake studies failed data quality control due to excessive artifacts in the recordings, leaving 25 sleep studies and 22 wake studies with analyzable device and biomarker data in the Villages Study. Of the participants enrolled in the UW Study, one was not compliant with the enforced wake protocol and was removed from analysis. A second participant was also excluded because of missing EEG powerband data during REM sleep caused by artifacts, which was required for the current analysis. The remaining 13 participants (55.9 ± 4.6 years of age; 6 female, 7 male) all completed the protocol. All overnight sleep data were usable, but two overnight wake studies were removed because of excessive artifact in the UW Study.

We measured plasma Aβ_40_, Aβ_42_, np-tau181, np-tau217, p-tau181, and p-tau217 at evening and morning timepoints in each participant prior to and following overnight sleep or sleep deprivation. Summary plasma AD biomarker levels are provided in **Table 3**. Of 38 participants, a total of 11 were assessed as ‘amyloid-positive’ by the C2N mass spectrometry with a Aβ_42_/ Aβ_40_ cutoff value of 0.089^32,34,35^ (**Table 2**). Plasma AD biomarker levels shown in **Table 3** are stratified by participant amyloid status. Overall, the measured plasma levels and overnight changes with sleep agree with prior reported *values* for Aβ_40_, Aβ_42_ and p-tau181^36^. Furthermore, the overnight changes in measured plasma levels following sleep and sleep-deprivation did not differ significantly, consistent with the possibility of competing clearance or production effects during sleep and sleep-deprivation.

**Table 3.** Morning and evening plasma AD biomarker levels in sleep and sleep deprivation conditions.

| Condition | Amyloid Status | Plasma Biomarker [units] | Morning Plasma Levels (*Conc<sub>AM</sub>*) | Evening Plasma Levels (*Conc<sub>PM</sub>*) | Differences (evening – morning) | DF | P (evening v morning) | P (sleep v wake) |
| --- | --- | --- | --- | --- | --- | --- | --- | --- |
| Sleep | Positive | Aβ<sub>40</sub> [pg/mL] | 401.8 (56.8) | 412.5 (72.6) | 10.71 (37.9) | 10 | 0.371 | 0.682 |
| | | Aβ<sub>42</sub> [pg/mL] | 32.2 (5.6) | 33.2 (6.2) | 1.05 (4.01) | 10 | 0.406 | 0.895 |
|  |  | np-tau181 [pg/mL] | 89.8 (20.8) | 92.7 (23.2) | 2.87 (10.9) | 10 | 0.401 | 0.118 |
|  |  | np-tau217 [pg/mL] | 101.3 (25.3) | 108.3 (25.7) | 7.05 (16.2) | 10 | 0.179 | 0.210 |
|  |  | p-tau181 [pg/mL] | 13.2 (4.7) | 14.4 (5.7) | 1.26 (1.7) | 10 | **0.035** | 0.102 |
| | Negative | Aβ<sub>40</sub> [pg/mL] | 411.6 (55.4) | 423.8 (50.5) | 12.18 (25.7) | 24 | **0.026** | 0.998 |
| | | Aβ<sub>42</sub> [pg/mL] | 42.4 (5.8) | 45.4 (5.8) | 2.94 (4.96) | 24 | **0.007** | 0.235 |
|  |  | np-tau181 [pg/mL] | 66.8 (19.3) | 68.5 (29.4) | 1.70 (15.5) | 26 | 0.574 | 0.218 |
|  |  | np-tau217 [pg/mL] | 83.2 (24.2) | 84.9 (35.3) | 1.68 (20.4) | 26 | 0.673 | 0.088 |
|  |  | p-tau181 [pg/mL] | 9.5 (3.0) | 10.2 (4.0) | 0.68 (2.22) | 26 | 0.122 | 0.791 |
| Sleep Deprivation / Wake | Positive | Aβ<sub>40</sub> [pg/mL] | 426.8 (69.8) | 431.4 (72.2) | 4.52 (28.6) | 8 | 0.648 | |
| | | Aβ<sub>42</sub> [pg/mL] | 35.1 (5.8) | 36.4 (5.4) | 1.26 (2.95) | 8 | 0.237 | |
|  |  | np-tau181 [pg/mL] | 84.7 (18.0) | 100.4 (34.9) | 15.64 (20.5) | 8 | 0.051 |  |
|  |  | np-tau217 [pg/mL] | 97.0 (23.0) | 116.2 (42.5) | 19.20 (23.6) | 8 | **0.040** |  |
|  |  | p-tau181 [pg/mL] | 13.8 (5.2) | 16.6 (6.3) | 2.80 (2.15) | 8 | **0.005** |  |
| | Negative | Aβ<sub>40</sub> [pg/mL] | 379.4 (35.2) | 391.6 (43.8) | 12.16 (33.7) | 22 | 0.098 | |
| | | Aβ<sub>42</sub> [pg/mL] | 39.6 (4.7) | 41.1 (5.5) | 1.56 (2.78) | 22 | **0.013** | |
|  |  | np-tau181 [pg/mL] | 57.1 (19.3) | 63.8 (25.6) | 6.69 (12.8) | 22 | **0.020** |  |
|  |  | np-tau217 [pg/mL] | 68.6 (21.4) | 79.2 (30.8) | 10.59 (15.7) | 22 | **0.004** |  |
|  |  | p-tau181 [pg/mL] | 8.1 (2.6) | 9.0 (3.0) | 0.85 (2.12) | 22 | 0.069 |  |

\*Data shown as mean (SD). *Conc<sub>AM</sub>*, morning plasma level; *Conc<sub>PM</sub>*, evening plasma level; DF, degrees of freedom; **Bolded P values reflect significant associations at P < 0.05 using a paired two-tailed Student's t-test of significance.** First column of P values (evening v morning) tests evening versus morning biomarker plasma level differences. Second column of P values (sleep v wake) tests sleep versus wake change in evening-morning differences of biomarker plasma levels.

We further investigated potential sequence-related effects within the randomized cross-over study design by comparing the evening-minus-morning plasma biomarker levels following sleep and/or sleep deprivation on the first visit with those on the second visit. This analysis was conducted separately for amyloid-positive and amyloid-negative groups, as well as for the combined amyloid positive and negative cohorts. A total of 40 distinct comparisons were made. Only two showed significant sequence differences between the first and second visits: (i) in the amyloid-negative group under sleep deprivation, **np-tau217** showed a difference (*P* = 0.031); (ii) in the combined amyloid group under sleep deprivation, **np-tau18** showed a difference (*P* = 0.043).

### Development of a multivariate model to relate release and glymphatic clearance features to plasma Aβ and tau concentrations

We developed a series of multivariate mixed models to define the effects that continuous features of glymphatic physiology and synaptic-metabolic activity have on overnight release and clearance of brain interstitial Aβ and tau to the plasma. Within these models, the multiple dependent variable measures for each participant were the morning plasma Aβ and tau (Aβ40, Aβ42, np-tau181, np-tau217, p-tau181) biomarker levels. Plasma p-tau217 levels were measured but excluded from analysis because a large proportion (9 out of 49) of these cognitively-intact individuals exhibited plasma p-tau217 levels below the limit of detection for the assay, consistent with prior study findings^37,38^. Data from amyloid-negative and amyloid-positive individuals were analyzed separately.

Because of the large number of measured outcomes, we used dimensionality reducing single-index regression^39^ to combine sleep-related factors into single ‘predictors’. A detailed explanation of this single predictor development is provided in **Supplementary Methods and Results**. Within these models, two distinct groups of predictors were analyzed: neurophysiological variables (***Physio***) and hypnographic sleep stages (***Hypno***). Separating predictors into these groups enabled comparison of both neurophysiological measures and sleep stage durations in explaining observed differences in morning plasma Aβ and tau levels, while mitigating multicollinearity among EEG power bands and sleep stages in the regression models, as previously documented^17^. The first group of predictors (***Physio***) included EEG non-rapid eye movement (NREM) delta (0.5–4 Hz) and theta power bands, REM sleep theta and beta power bands, heart rate variability (HRV), and pulse transit time (PTT) during NREM, and parenchymal resistance R_P_. HRV and PTT values during REM sleep were highly correlated with those during NREM and were excluded. The NREM and REM power bands selected represent the majority of the spectral power during these sleep stages, thus contributing to synaptic-metabolic release^30,31^, and because EEG delta, beta and theta power bands have established associations with glymphatic clearance^13,17,22,23^. The second group (***Hypno***) comprised hypnographic sleep stages, including the durations of REM, N1, NREM (N2 + N3), and wake after sleep onset (WASO). Each group of individual predictors was combined using linear combinations into three single-index predictors for amyloid-negative and amyloid-positive participants separately: the neurophysiological predictor under the sleeping condition (***Physio***_***S***_) and under the sleep deprivation/wake condition (***Physio***_***W***_), and the sleep hypnogram predictor under the sleeping condition (***Hypno***_***S***_). The optimal predictor components and weights for the sleep condition (***Physio***_***S***_, ***Hypno***_***S***_) and sleep deprivation/wake condition (***Physio***_***W***_), where the sum of the squared weights normalizes to unity, are provided in **Table 4**.

**Table 4.** Full model predictors and optimal weights predicting morning plasma AD biomarker levels in sleep and sleep deprivation conditions.

| Condition and Predictor | Amyloid Status | Predictors | Estimate | CI | Wald's P |
| --- | --- | --- | --- | --- | --- |
| Sleep Condition<br>*Physios* | Positive | **R<sub>p</sub>** | **0.968** | **0.177 – 0.998** | **0.036** |
|  |  | PTT (NREM) | -0.034 | -0.396 – 0.132 | 0.388 |
|  |  | **HRV (NREM)** | **-0.068** | **-0.464 – -0.014** | **0.030** |
|  |  | EEG Delta Power (NREM) | 0.086 | -0.198 – 0.705 | 0.399 |
|  |  | EEG Theta Power (NREM) | 0.162 | -0.261 – 0.604 | 0.195 |
|  |  | EEG Theta Power (REM) | -0.146 | -0.547 – 0.496 | 0.436 |
|  |  | EEG Beta Power (REM) | -0.041 | -0.671 – 0.095 | 0.264 |
|  | Negative | **R<sub>p</sub>** | **0.963** | **0.948 – 0.977** | **<0.001** |
|  |  | **PTT (NREM)** | **-0.058** | **-0.200 – -0.028** | **0.006** |
|  |  | HRV (NREM) | 0.114 | -0.003 – 0.178 | 0.060 |
|  |  | **EEG Delta Power (NREM)** | **-0.041** | **-0.149 – -0.010** | **0.026** |
|  |  | **EEG Theta Power (NREM)** | **0.169** | **0.083 – 0.218** | **<0.001** |
|  |  | **EEG Theta Power (REM)** | **0.087** | **0.001 – 0.133** | **0.058** |
|  |  | **EEG Beta Power (REM)** | **-0.135** | **-0.250 – -0.093** | **<0.001** |
| Sleep Condition<br>*Hypnos* | Positive | NREM Duration | -0.182 | -0.629 – 0.108 | 0.166 |
|  |  | REM Duration | -0.519 | -0.829 – 0.337 | 0.209 |
|  |  | **N1 Duration** | **0.834** | **0.366 – 0.990** | **0.010** |
|  |  | WASO Duration | 0.044 | -0.396 – 0.477 | 0.620 |
|  | Negative | **NREM Duration** | **-0.090** | **-0.239 – -0.054** | **0.001** |
|  |  | **REM Duration** | **0.525** | **0.423 – 0.585** | **<0.001** |
|  |  | **N1 Duration** | **0.801** | **0.754 – 0.855** | **<0.001** |
|  |  | **WASO Duration** | **0.273** | **0.131 – 0.344** | **0.019** |
| Sleep Deprivation/Wake<br>*Physio<sub>w</sub>* | Positive | R<sub>p</sub> | 0.621 | -0.365 – 0.891 | 0.087 |
|  |  | PTT | 0.190 | -0.134 – 0.505 | 0.390 |
|  |  | HRV | -0.242 | -0.450 – 0.106 | 0.166 |
|  |  | **EEG Delta Power** | **-0.487** | **-0.831 – -0.124** | **0.027** |
|  |  | EEG Theta Power | -0.382 | -0.670 – 0.584 | 0.339 |
|  |  | EEG Beta Power | 0.371 | -0.036 – 0.690 | 0.071 |
|  | Negative | **R<sub>p</sub>** | **0.626** | **0.519 – 0.651** | **<0.001** |
|  |  | PTT | -0.004 | -0.127 – 0.029 | 0.461 |
|  |  | **HRV** | **0.136** | **0.058 – 0.173** | **0.028** |
|  |  | **EEG Delta Power** | **-0.658** | **-0.717 – -0.634** | **<0.001** |
|  |  | **EEG Theta Power** | **-0.391** | **-0.473 – -0.359** | **<0.001** |
|  |  | EEG Beta Power | 0.062 | -0.026 – 0.095 | 0.170 |

\*CI, 95% Confidence Interval; R<sub>p</sub>, Parenchymal Resistance; PTT, Pulse Transit Time; HRV, Heart Rate Variability; EEG, Electroencephalography; ***Physios*** and ***Physio<sub>w</sub>***, single predictors constructed from the linear combination of the full model predictors during the sleep and sleep deprivation (wake) conditions, respectively, for both amyloid positive and negative participants separately; ***Hypnos***, single predictor constructed from the full model hypnogram features during the sleep condition for both amyloid positive and negative participants separately. The estimated weights were selected using optimization to maximize the log likelihood of the production/clearance model. **Bolded P values reflect significant associations at P < 0.05 using bias-corrected bootstrapping.**

We developed parallel multivariate linear mixed models representing the **null model** and the **neuro-glymphatic model**, as described in detail in **Supplemental Methods and Results**. Under the **null model**, morning plasma levels of Aβ and tau species were not dependent upon sleep-active glymphatic exchange and synaptic-metabolic release, and thus were not assumed to be influenced by sleep neurophysiological (***Physio***) and sleep stage (***Hypno***) predictors. Rather the morning plasma levels of AD biomarkers (***Conc***_***AM***_) were regressed on the evening plasma levels (***Conc***_***PM***_) for Aβ and tau biomarkers, separately for amyloid positive and amyloid negative participants. Potentially confounding variables age, sex, APOE-ε4 status, and study site were included in the model. A circadian confounder was also included in the model, reflecting the interval between the evening AD biomarker sample time and sleep-onset measured by EEG. Multivariate mixed models used participant ID as a random intercept and the categorical *biomarker* variable as a vector of random slopes. The **neuro-glymphatic model** shares the features of the **null model** but included the effects of the single index predictors (***Physio***_***S***_, ***Physio***_***W***_ or ***Hypno***_***S***_), and their respective interaction terms with evening levels of plasma AD biomarkers (***Physio***_***S***_ * ***Conc***_***PM***_, ***Hypno***_***S***_ ***\* Conc***_***PM***_, ***Physio***_***W***_ ***\* Conc***_***PM***_). The likelihood ratio test (LRT) of the **neuro-glymphatic model** versus the **null model** was used to determine which model performed better at predicting the morning plasma Aβ and tau biomarker levels. The conditional variances of each model, i.e. the residual variation *conditional on* a participant’s random effects, were used to define the amount of variance in morning plasma Aβ and tau levels explained by the **neuro-glymphatic model** over that explained by the **null model**. This is described in detail in **Supplementary Methods and Results**.

### Effects of sleep-related neurophysiological features on overnight Aβ and tau release and clearance

The predictor ***Physio***_***S***_ that led to the best fit of the data based on maximum likelihood for the **neuro-glymphatic model** in sleep for both amyloid-negative and -positive individuals showed that R_P_ was the overwhelming contributor to ***Physio***_***S***_ (**Table 4**). The output of the **null model** and **neuro-glymphatic model** with the predictor ***Physio***_***S***_ for both amyloid-positive and amyloid-negative individuals are shown in **Table 5**. Estimates for single predictor coefficients within the **neuro-glymphatic model** for each plasma AD biomarker, evaluated at the mean evening biomarker level, are provided in **Supplemental Table 7**.

**Table 5:**
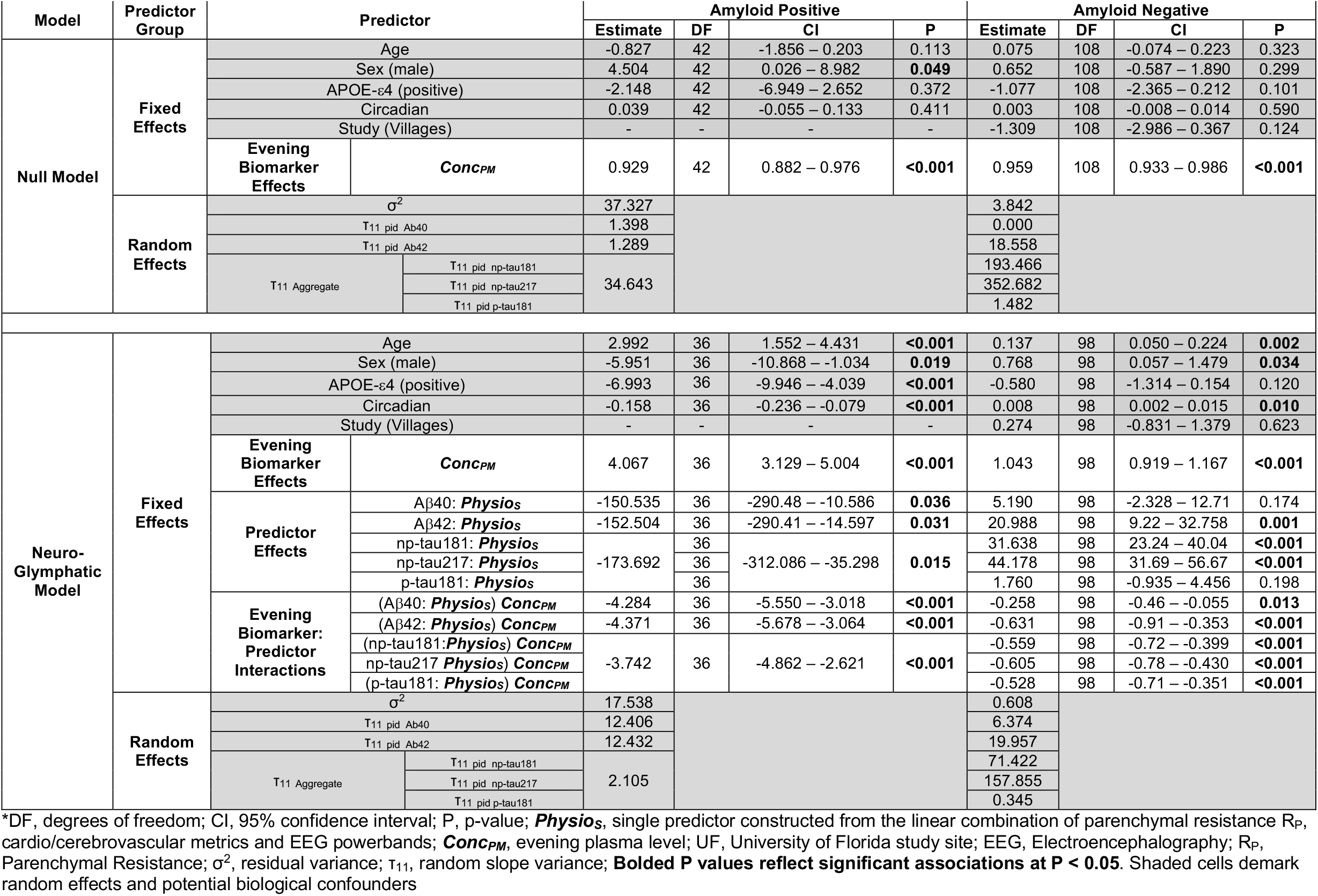
Prediction of morning plasma AD biomarker levels following overnight sleep using the *null model* and the *neuro-glymphatic model* with *Physio*_*S*_.

Within the **null model**, evening plasma Aβ and tau levels (***Conc***_***PM***_) were the main predictor of morning plasma levels in both amyloid-positive and -negative participants, reflecting between-participant evening consistency in AD biomarker levels (**Table 5**). Within the **neuro-glymphatic model**, in addition to evening plasma biomarker levels, participant age, sex, and circadian alignment each significantly contributed to morning plasma Aβ and tau levels in both amyloid-positive and -negative participants. APOE-e4 status was significantly associated with lower morning AD biomarker levels in amyloid-positive, but not amyloid-negative individuals. Age was significantly associated with higher morning AD biomarker levels in both groups. Across both amyloid-positive and -negative participants, the ***Physio***_***S***_ predictor significantly impacted plasma Aβ and tau levels, and the ***Conc***_***PM***_\****Physio***_***S***_ interaction terms were consistently significant across Aβ and tau analytes (**Table 5**).

By including overnight R_P_, PTT, HRV, and EEG power bands in the single predictor ***Physio***_***S***_, the **neuro-glymphatic model** led to greater predictive performance over the **null model** (**Table 6**, LRT, p <0.001). When comparing the percent variance of morning plasma AD biomarker levels explained by the **neuro-glymphatic model** compared to the **null model**, inclusion of overnight ***Physio***_***S***_ in the **neuro-glymphatic model** explained between 49.1% (Aβ40) to 56.0% (Aβ42) additional variance in amyloid-positive individuals, and between 71.3% (p-tau181) to 97.8% (Aβ40) in amyloid-negative individuals (**Table 7**).

**Table 6:** Likelihood ratio test of the *neuro-glymphatic model* versus the *null model*\*.

| Condition and Predictor | Amyloid Status | Model | AIC | BIC | logLik | LRT | DF | P |
| --- | --- | --- | --- | --- | --- | --- | --- | --- |
| Sleep Condition, *Physio<sub>s</sub>* | Positive | Null | 396.0 | 422.1 | -185.0 |  |  |  |
|  |  | Model | 362.9 | 401.0 | -162.4 | 45.2 | 6 | **<0.001** |
|  | Negative | Null | 814.5 | 880.6 | -384.2 |  |  |  |
|  |  | Model | 735.4 | 830.2 | -334.7 | 99.1 | 10 | **<0.001** |
| Sleep Condition, *Hypno<sub>s</sub>* | Positive | Null | 396.0 | 422.1 | -185.0 |  |  |  |
|  |  | Model | 381.4 | 419.5 | -171.7 | 26.6 | 6 | **<0.001** |
|  | Negative | Null | 814.5 | 880.6 | -384.2 |  |  |  |
|  |  | Model | 741.6 | 836.5 | -337.8 | 92.9 | 10 | **<0.001** |
| Sleep Deprivation/Wake *Physio<sub>w</sub>* | Positive | Null | 337.4 | 360.9 | -155.7 |  |  |  |
|  |  | Model | 281.0 | 315.3 | -121.5 | 68.4 | 6 | **<0.001** |
|  | Negative | Null | 688.6 | 751.8 | -321.3 |  |  |  |
|  |  | Model | 623.5 | 714.1 | -278.8 | 85.1 | 10 | **<0.001** |

\*AIC, Akaike Information Criteria; BIC, Bayesian Information Criteria; logLik, log likelihood; LRT, log likelihood ratio; DF, degrees of freedom; P, p-value; ***Physio<sub>s</sub>*** and ***Physio<sub>w</sub>***, single predictors constructed from the linear combination of the full model predictors during the sleep and sleep deprivation (wake) conditions, respectively, for both amyloid positive and negative participants separately; ***Hypno<sub>s</sub>***, single predictor constructed from the full model hypnogram features during the sleep condition for both amyloid positive and negative participants separately; **Bolded P values reflect significant LRT at P < 0.05.**

**Table 7.** Percent of variance in morning plasma AD biomarker levels explained by the *neuro-glymphatic model* over the *null model*\*.

| Condition and Predictor | Amyloid Status | Plasma Biomarker | Variance Explained | CI |
| --- | --- | --- | --- | --- |
| Sleep Condition *Physio<sub>s</sub>* | Positive | All Biomarkers | 51.9% | 38.6% – 84.3% |
|  |  | Ab40 | 49.1% | 22.4% – 92.8% |
|  |  | Ab42 | 56.0% | 5.4% – 91.4% |
|  |  | All Tau | 51.7% | 35.6% – 83.7% |
|  | Negative | All Biomarkers | 91.1% | 47.4% – 98.1% |
|  |  | Ab40 | 97.8% | 45.5% – 99.6% |
|  |  | Ab42 | 93.7% | 40.7% – 99.3% |
|  |  | np-tau217 | 95.4% | 42.9% – 99.3% |
|  |  | np-tau181 | 94.1% | 38.7% – 99.1% |
|  |  | p-tau181 | 71.3% | 49.0% – 97.3% |
| Sleep Condition *Hypno<sub>s</sub>* | Positive | All Biomarkers | 42.0% | 32.5% – 81.8% |
|  |  | Ab40 | 57.7% | 16.1% – 91.4% |
|  |  | Ab42 | 62.5% | 9.1% – 90.8% |
|  |  | All Tau | 38.4% | 28.9% – 80.7% |
|  | Negative | All Biomarkers | 87.4% | 17.7% – 96.9% |
|  |  | Ab40 | 95.7% | -30.1% – 98.3% |
|  |  | Ab42 | 87.1% | -39.7% – 97.2% |
|  |  | np-tau217 | 93.1% | 21.4% – 98.6% |
|  |  | np-tau181 | 90.9% | 1.5% – 98.4% |
|  |  | p-tau181 | 64.0% | 26.4% – 96.8% |
| Sleep Deprivation/Wake *Physio<sub>w</sub>* | Positive | All Biomarkers | 68.9% | 65.5% – 92.5% |
|  |  | Ab40 | 76.6% | -7.9% – 94.1% |
|  |  | Ab42 | -53.8% | -13.7% – 95.4% |
|  |  | All Tau | 72.8% | 68.0% – 92.7% |
|  | Negative | All Biomarkers | 85.1% | 34.9% – 98.0% |
|  |  | Ab40 | 92.5% | 40.2% – 99.4% |
|  |  | Ab42 | 93.3% | 44.7% – 99.5% |
|  |  | np-tau217 | 60.7% | -184.8% – 95.9% |
|  |  | np-tau181 | 84.3% | -44.6% – 98.7% |
|  |  | p-tau181 | 73.8% | 34.9% – 96.8% |

\*CI, 95% confidence interval; P, p-value; ***Physio<sub>s</sub>*** and ***Physio<sub>w</sub>***, single predictors constructed from the linear combination of the full model predictors during the sleep and sleep deprivation (wake) conditions, respectively, for both amyloid positive and negative participants separately; ***Hypno<sub>s</sub>***, single predictor constructed from the full model hypnogram features during the sleep condition for both amyloid positive and negative participants separately. **Bolded P values reflect significant values at P < 0.05.**

As detailed above, the ***Physio***_***S***_ single-index predictor includes several sleep-related physiological features that could contribute variously to changes in glymphatic clearance to the plasma, or to synaptic-metabolic release of Aβ and tau. We carried out a sensitivity analysis to explore the relative contributions of these individual neurophysiological predictors, and of glymphatic clearance versus synaptic-metabolic release to changes in morning plasma Aβ and tau analyte levels. This sensitivity analysis is provided in detail in **Supplementary Methods and Results**. The analysis suggests that parenchymal resistance R_P_, cerebrovascular compliance (measured by PTT) and NREM EEG delta power during sleep, which are each associated with enhanced glymphatic exchange in rodents and humans^12,13,17,22,23,25,40,41^, facilitated the overnight clearance of brain interstitial Aβ and tau into the plasma made the largest contribution to changes in plasma Aβ and tau levels. Relative to the contributions of these individual predictors, the contribution from increased synaptic-metabolic activity reflected in NREM EEG theta power and REM EEG theta and beta power were comparatively minor (**Supplementary Table 3**). The combined effects of these predictors which impact both glymphatic clearance and synaptic-metabolic release of Aβ and tau accounted for over 50% of the variance in morning plasma Aβ and tau levels in amyloid-positive individuals, and over 90% of the variance in amyloid-negative individuals, compared to that explained by the **null model** alone (**Table 7**). These data suggest that both increased synaptic-metabolic release and glymphatic clearance of Aβ and tau during sleep contribute to increased morning plasma AD biomarker levels, but that increased clearance driven by reduced R_P_, increased cerebrovascular compliance, and increased EEG delta power are the greatest contributors to these effects.

### NREM sleep duration enhanced overnight clearance of Aβ and tau to the plasma

We next tested whether differences in overnight sleep stages influenced the glymphatic clearance or synaptic-metabolic release of Aβ and tau to the plasma. The predictor ***Hypno***_***S***_ was constructed from regressors chosen from EEG hypnogram stages REM, N1, NREM (N2, N3) and WASO that led to the best fit of the data based on the maximum likelihood optimization for the **neuro-glymphatic model** for amyloid-positive and amyloid-negative individuals (**Table 4**). The **null model** and the **neuro-glymphatic model** fitted to the morning plasma AD biomarker level data following overnight sleep with the ***Hypno***_***S***_ predictor are shown in **Supplemental Table 10**. By including overnight REM, NREM, N1 and WASO in the single predictor ***Hypno***_***S***_, the **neuro-glymphatic model** led to greater predictive performance over the **null model** (**Table 6**, LRT, p <0.001). When comparing the percent variance of morning plasma AD biomarker levels that was explained by overnight sleep stage durations in the **neuro-glymphatic model** and that was not explained by the **null model**, the **neuro-glymphatic model** in amyloid-positive individuals explained between 38% (all tau species) to 62.5% (Aβ42) of the variance (**Table 7**). In amyloid-negative individuals, sleep stage duration did not perform better than the **null model** in explaining Aβ40 and Aβ42 levels, but explained between 64% (p-tau181) and 93.1% (np-tau217) of the tau variance (**Table 7**).

Using a similar approach to that above, we observed that at a mean value of each evening plasma AD biomarker level, a decrease in overnight ***Hypno***_***s***_, which corresponds to an increase in NREM sleep duration and a decrease in REM sleep, N1 sleep, and WASO duration (**Table 4**), increases morning plasma Aβ and tau biomarker levels (**Supplemental Table 7**). Sensitivity analysis, as described in **Supplementary Methods and Results** (**Supplementary Table 3–4**), suggests that increased NREM sleep duration, which is associated with increased glymphatic function in rodents and humans^13,16,17^, increases the clearance of Aβ and tau to the plasma. A decrease REM sleep, N1 sleep and WASO duration were also associated with an increase in glymphatic clearance (**Supplementary Table 3**), primarily because of their negative correlation with NREM sleep duration. These effects explain approximately 50% of the unexplained variance in the morning plasma AD biomarker levels from the **null model** in amyloid-positive individuals but fail to explain additional Aβ40 and Aβ42 variance in amyloid-negative individuals (**Table 7**). These data demonstrate that increased NREM (N2 + N3) sleep duration contributes to the overnight clearance of Aβ and tau to the plasma.

### Combined effects of release and clearance to plasma Aβ and tau levels during sleep deprivation

We next evaluated whether the neurophysiological features used to characterize sleep also influenced Aβ and tau release and clearance under conditions of overnight sleep deprivation (waking). To address this, we constructed the single-index predictor ***Physio***_***W***_ from the same regressors, optimizing it to best fit the sleep deprivation data (**Table 4**). The **null model** and the **neuro-glymphatic model** fitted to morning plasma AD biomarker levels following sleep deprivation, using the single predictor ***Physio***_***W***_ are shown **Supplemental Table 11**. By incorporating overnight neurophysiological features related to glymphatic exchange and synaptic-metabolic activity into ***Physio***_***W***_, the **neuro-glymphatic model** demonstrated superior predictive performance for morning plasma AD biomarker levels compared with the null model (**Table 6**, LRT, p < 0.001). When assessing the additional variance in morning plasma Aβ and tau levels explained by the **neuro-glymphatic model** beyond that explained by the **null model**, we found that in amyloid-positive individuals the model accounted for 72.8% of all tau variance. In amyloid-negative individuals, the model explained 92.5% of Aβ40, 93.3% of Aβ42, and 73.8% of p-tau181 variance (**Table 7**).

In individuals undergoing overnight sleep deprivation, HRV, PTT, and R_P_ measures contributed to Aβ and tau release and clearance in a manner largely consistent with their effects during sleep, although the contribution of R_P_ was substantially smaller (**Supplementary Table 3**). In this context, increases in EEG delta and theta power were associated with elevated Aβ and tau release, whereas a reduction in EEG beta power enhanced clearance, an association previously observed in both rodent and human studies^17,22^. Note that in amyloid-positive individuals, predictor assignment for a decrease in ***Physio***_***W***_ was based on all tau effects (**Supplemental Table 8**), as Aβ40 and Aβ42 effects were not statistically significant (**Supplemental Table 11**).

These data suggest that under conditions of sleep deprivation, both synaptic-metabolic release and glymphatic clearance of Aβ and tau contribute to morning plasma AD biomarker levels.

### The predictors Physio_S_, Hypno_S_ and Physio_W_ in the neuro-glymphatic model replicate the rate constants in the compartmental pharmacokinetic model

The linear interaction terms between the predictors ***Physio***_***S***_, ***Hypno***_***S***_ and ***Physio***_***W***_ and evening plasma AD biomarker levels in the three **neuro-glymphatic models** for amyloid-positive and amyloid-negative individuals were significant at both the group level (**Table 8**) and individual levels (**Table 5, Supplemental Tables 5–6**). These interaction terms are the same as described by **Equation 3** in the solution of the multi-compartment model (**Supplementary Methods and Results**). That is, the linear interaction terms between the predictors ***Physio***_***S***_, ***Hypno***_***S***_ and ***Physio***_***W***_ and evening plasma AD biomarker levels from experimental observations are consistent with those derived from multicompartment first-order kinetic modeling under the **neuro-glymphatic model** assumption with ISF to CSF clearance rate constants and synaptic-metabolic release that are time-varying through ***Physio***_***S***_, ***Hypno***_***S***_ and ***Physio***_***W***_.

**Table 8.**
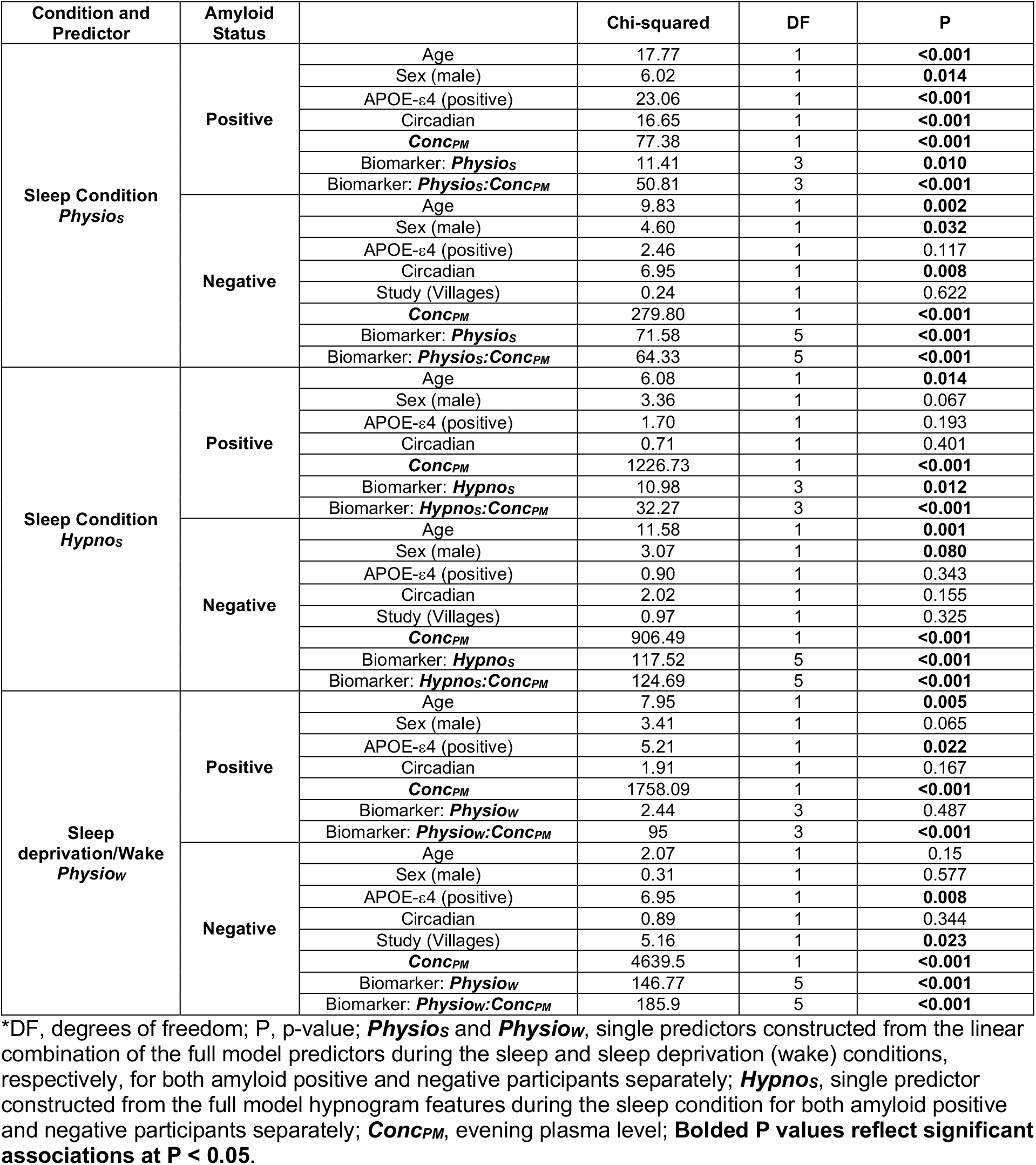
Analysis of deviance using Type III Wald Chi-square test for *neuro-glymphatic model* interaction effects*.

To further demonstrate this correspondence, we compared experimental predictions of morning plasma AD biomarker levels when ***Physio***_***S***_ and ***Hypno***_***S***_ were varied by ±1 standard deviation with predictions generated by the compartmental model under proportional variation of the rate constants *k*_*glymph*_*out*_, *k*_*glymph*_*in*_ and *k*_*cell*_*rel*_. As shown in **Figure 3A–B**, varying the magnitude of fitted predictors in the **neuro-glymphatic models** for sleep, while proportionally varying *k*_*glymph*_*out*_ and *k*_*glymph*_*in*_ in the compartment model, produced similar changes in predicted morning plasma AD biomarker levels across all conditions. Furthermore, the compartmental model reproduced the experimentally-observed pattern of greater clearance of aggregation-prone species (Aβ42, p-tau181) relative to non-aggregation-prone species (Aβ40, np-tau181), consistent with an increased Aβ42/Aβ40 and p-tau181/np-tau181 ratio over baseline clearance. By contrast, increasing *k*_*cell*_*rel*_ in the compartmental model generated similar directional effects on morning plasma biomarker levels but reduced clearance of aggregation-prone species relative to non-aggregation-prone species, a pattern inconsistent with the experimental predictions (**Figure 1, Supplemental Table 1**). Taken together, these findings support glymphatic clearance as the predominant physiological mechanism underlying the observed variation in morning plasma AD biomarker levels following overnight sleep.

**Figure 3.**
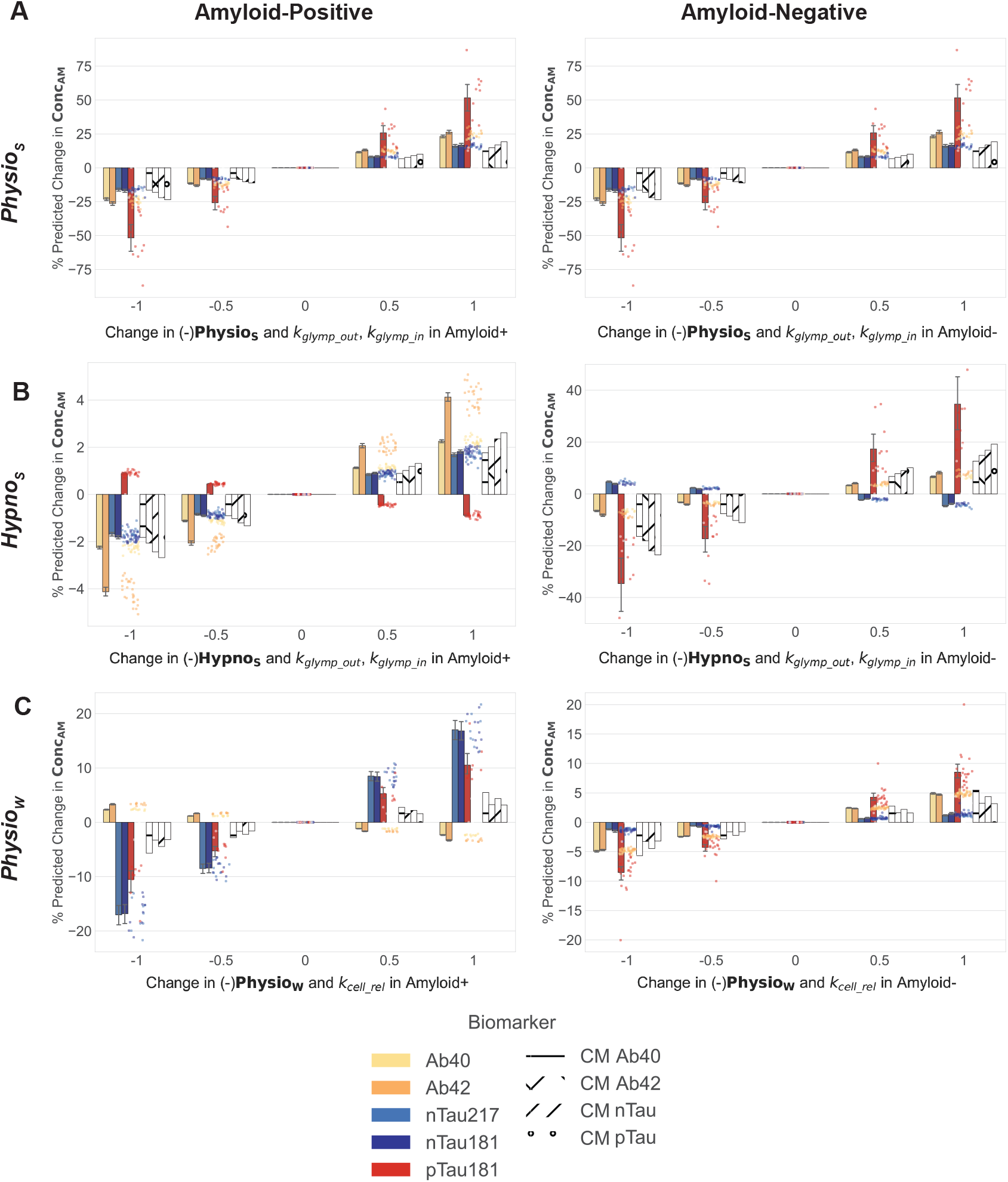
Comparison of neuro-glymphatic model and multicompartment model predictions of morning plasma level. The percent change prediction in morning plasma level ***Conc***_***AM***_ of AD biomarkers Aβ_40_, Aβ_42_, np-tau181, np-tau217, p-tau181 resulting from a change in units of SD of the single predictors ***Physio***_***S***_ **(A), *Hypno***_***S***_ **(B)**, and ***Physio***_***W***_ **(C)** of the **neuro-glymphatic model** in amyloid-positive and amyloid-negative participants are compared against the percent change in the morning amyloid β and tau multicompartment model prediction from a corresponding change glymphatic efflux/influx rate constants *k*_*glymph*_*out*_ and *k*_*glymph*_*in*_ **(A)** and **(B)**, or synaptic and metabolic release rate constant *k*_*cell*_*rel*_ **(C)**. Predicted mean and 95% confidence intervals for Aβ_40_, Aβ_42_, np-tau181, np-tau217, p-tau181 at each SD change in the single predictor are shown in order as the first five bars alongside the **neuro-glymphatic model** predictions for that SD change. Multicompartment model predictions for Aβ40, Aβ42, non-phosphorylated and phosphorylated tau at each corresponding change of the rate constants *k*_*glymph*_*out*_ and *k*_*glymph*_*in*_ **(A)** and **(B)**, or *k*_*cell*_*rel*_ **(C)**, are shown in order as the last four hatched bars. The change in the negative of the single predictors are shown for comparison with the change in the rate constants. For example, a 1SD increase in the negative predictor corresponds to a 1SD decrease in the actual predictor. (2025). https://github.com/appliedcognition/ac_compartment_model_simulation

We next compared experimental predictions of morning plasma AD biomarker levels during sleep deprivation, obtained by varying ***Physio***_***W***_ by ±1 standard deviation, with predictions from the compartment model under proportional variation of the rate constants *k*_*glymph*_*out*_, *k*_*glymph*_*in*_ and *k*_*cell*_*rel*_. As shown in **Figure 3C**, reproducing the experimental results from the **neuro-glymphatic models** required varying *k*_*cell*_*rel*_ in the compartmental model. This adjustment produced similar changes in predicted morning plasma AD biomarker levels and replicated the experimentally-observed pattern of reduced clearance of aggregation-prone species (Aβ42, p-tau181) relative to non-aggregation-prone species (Aβ40, np-tau181). Thus, under sleep deprivation, increased synaptic-metabolic release emerges as the predominant physiological mechanism underlying the observed morning plasma AD biomarker levels.

It is noteworthy that while the results of the statistical and compartmental models converge, they were derived from independent sources. The glymphatic models were statistically estimated from the experimental data from the Villages and UW clinical studies whereas the compartmental model was based on the elimination kinetics of amyloid β and tau obtained from independent published research^27,28^.

## DISCUSSION

We developed a compartmental transport model based on published kinetic values to predict the effect that sleep-active glymphatic CSF-ISF exchange and synaptic-metabolic activity has on overnight changes in plasma Aβ and tau levels. To rigorously test the glymphatic model of interstitial Aβ and tau clearance from the human brain during sleep, and to define the relative contributions that changes in synaptic-metabolic release and clearance make to plasma Aβ and tau levels, we conducted a multi-site, randomized, crossover clinical study. This study utilized a novel investigational device from Applied Cognition^17^ to measure parenchymal resistance R_P_, sleep EEG features, cerebrovascular pulse transit time PTT, heart rate variability HRV, in addition to clinically validated blood-based biomarkers of Aβ and tau provided by C2N^32–34^. Healthy older participants underwent both overnight sleep and sleep deprivation visits. Our findings showed that reduced parenchymal resistance, increased cerebrovascular compliance, and elevated EEG delta power during NREM sleep – each of which are associated with increased glymphatic function in rodents and humans^12,13,17,22,23,25,40^ - predicted higher morning plasma levels of Aβ and tau. EEG theta power during NREM sleep, and both theta and beta power during REM sleep, also contributed to morning plasma Aβ and tau levels, though to a lesser extent, suggesting a role for synaptic-metabolic activity in Aβ and tau release to the plasma^29–31^. These predictors of sleep-active glymphatic clearance and synaptic-metabolic release explained over 50% of the residual variance in morning plasma AD biomarker levels among amyloid-positive individuals, and over 90% among amyloid-negative individuals, that was not explained by the model that assumed constant overnight CSF–ISF exchange and reduced but uniform synaptic–metabolic activity across individuals.

The burden of Aβ and phosphorylated tau in the brain is diagnostic for AD and prognostic of its progression^42–44^. Recently approved monoclonal antibody therapies targeting Aβ demonstrate marked reduction in brain Aβ burden, but at present provide only modest improvement in cognitive outcomes and quality of life^45,46^. This apparent shortcoming has led to an increasing interest in extending anti-Aβ immunotherapy into earlier, preclinical stages of AD and into at-risk populations such as *APOE-ε4* carriers. It is also possible that if the mechanisms contributing to the development and progression of Aβ and tau pathology can be defined, then it may be possible both to better identify individuals at risk of the development of this pathology and to intervene in at-risk individuals to prevent its development and eventual progression.

In rodent models, sleep-active glymphatic function has been implicated in the clearance of both Aβ and tau. Interstitial Aβ and tau both move through brain tissue along perivascular pathways^10,14^, while inhibition of glymphatic function by either *Aqp4* or *Snta1* gene deletion slows the clearance of Aβ and tau and promotes the development of Aβ and tau pathology^10,13–15,47–51^. Glymphatic function is impaired in animal models of aging^52^, cerebrovascular dysfunction^53,54^, traumatic brain injury^14,55–57^, and sleep disruption^13,58^; each of which are non-genetic risk factors for Alzheimer’s disease. This body of results from rodent studies suggests that impairment of glymphatic clearance of Aβ and tau is a key factor in the development of AD, serving as a mechanistic linkage between a wide spectrum non-genetic AD risk factors and the development of AD-related Aβ and tau pathology^59,60^. If true, then the detection of glymphatic impairment would permit the identification of individuals at risk for the development of Aβ and tau pathology, while targeting glymphatic function would provide a novel approach to the prevention of the development and progression of AD pathology in at-risk individuals.

Despite promising results from rodent studies, key biological features of sleep-active glymphatic function have only recently been confirmed in the human brain. Intrathecal contrast-enhanced MRI studies have demonstrated extensive CSF-ISF exchange in the human brain^61^, that this exchange is organized along the axis of the cerebral arterial vasculature^62^, and that ISF solute clearance is more rapid in the sleeping compared to the waking brian^16^. In a recent study, we demonstrated that glymphatic function is sleep-active, and enhanced during sleep by increasing EEG delta power, reduced EEG beta power, reduced HR, and reduced parenchymal resistance R_P_^17^. Yet a role for sleep active glymphatic exchange in the clearance of Aβ and tau from the human brain remains largely undefined.

One recent study by Lucey and colleagues reported that the CSF-to-blood clearance of Aβ and tau are impaired by acute sleep deprivation^4^, although this study did not specifically implicate glymphatic clearance in this association. In another important study by Eide et al.^21^, the authors report that in participants undergoing assessment of glymphatic function and CSF-to-plasma clearance by intrathecal contrast-enhanced MRI, slowed glymphatic clearance was associated with lower plasma total-tau levels, while slowed CSF-to-plasma clearance was correlated with lower Aβ42 levels. While this study supports a role for glymphatic exchange in the clearance of AD-related biomarkers, the interpretation of the findings has key limitations. Because intrathecal administration is an off-label use of gadolinium-based contrast agents such as gadobutrol, the study was carried out in participants being evaluated for disordered CSF circulation and not otherwise ‘healthy’ participants. This introduced a several decade age range and multiple clinical indications in the study group that would be expected to confound the relationship between plasma levels of AD biomarkers and glymphatic function. While the study did evaluate the effect of time of day on plasma biomarker levels, the relationship between measures of glymphatic clearance and overnight dynamic changes in AD biomarker levels, rather than ‘steady state’ levels, was not evaluated. Lastly, while overall self-reported sleep quality was evaluated with the Pittsburgh Sleep Quality Index, objective measures of sleep parameters, sleep stages, or sleep EEG spectral band powers were not evaluated. In the present study, we extend these initial studies by evaluating the effect of overnight parenchymal resistance, sleep stages, sleep EEG powerbands during REM and NREM, cerebrovascular arterial compliance and heart-rate variability, each of which is a key determinate of glymphatic transport^12,13,17,22,23,25,40,41^ or a key contributor to synaptic-metabolic release^29–31^, on the dynamic clearance of Aβ and tau to the plasma in healthy older individuals.

To test whether sleep-active glymphatic function contributes to the clearance of Aβ and tau from the human brain independent of variations in synaptic-metabolic activity during sleep, we first developed a multicompartment model of Aβ and tau clearance from the brain interstitium. The model was designed to predict the effects of sleep-related changes in glymphatic CSF-ISF exchange and synaptic-metabolic release on evening-to-morning changes plasma Aβ and tau levels. Glymphatic clearance was modeled as changes in the rate constants between ISF and CSF compartments, demonstrating that an overnight increase in exchange would result in higher plasma AD biomarker levels the following morning. Similarly, synaptic-metabolic release was modeled as a change in the rate constant for Aβ and tau release from cells into the ISF, which also predicted elevated morning plasma biomarker levels when release increased.

Monomeric Aβ42 and p-tau181 in the ISF were modeled as being in equilibrium with their non-monomeric forms using a separate compartment and published rate constants between ISF and aggregate pools^27,28^. This revealed that an increase (or decrease) in the exchange rate between ISF and CSF (glymphatic exchange) preferentially increased (or decreased) the clearance of aggregation-prone species (Aβ42 and p-tau181) relative to non-aggregation-prone species (Aβ40 and np-tau181). In contrast, increasing the release rate from cells into the ISF led to the opposite effect, preferentially elevating non-aggregation-prone over aggregation-prone species. These key differences in model behavior allowed us to directly test the hypothesis that glymphatic clearance of Aβ and tau is the dominant contributor to sleep-related variation in plasma Aβ and tau levels, while synaptic–metabolic release dominates during sleep deprivation (and likely waking).

Analyzing data from two cross-over design clinical studies, we evaluated the validity of two parallel models to explain overnight changes in plasma Aβ and tau levels. Our **null hypothesis** was that neither glymphatic exchange nor synaptic-metabolic release contributed to the overnight changes in AD plasma biomarkers, and the **null model** used only the evening AD plasma biomarker levels adjusted by biological confounders to predict the morning AD biomarker levels. The **neuro-glymphatic hypothesis** was that increased CSF-ISF exchange during sleep increased the overnight clearance of Aβ and tau from brain to the plasma and that individual differences in overnight synaptic-metabolic release further altered morning plasma levels. Because glymphatic function and synaptic-metabolic activity during sleep are regulated by changes in EEG delta, theta and beta power during NREM and REM, and PTT, HRV, and R_P_^12,13,17,22,23,25,29–31,40,41^, we tested if the inclusion of these features would improve the prediction of plasma Aβ and tau levels consistent with our multicompartment model prediction.

Using a single-index regression that linearly combined these variables into single predictor ***Physio***_***S***_ for sleep, the **neuro-glymphatic model** explained over 50% of the residual variance in morning plasma AD biomarker levels in amyloid-positive individuals and over 90% in amyloid-negative individuals during the sleep condition. The optimal sleep predictor ***Physio***_***S***_ which maximized the log-likelihood during sleep, was primarily driven by parenchymal resistance R_P_, with smaller contributions from cerebrovascular compliance measure by PTT and EEG delta power during NREM sleep, features associated with increased glymphatic clearance. Minor contributions also came from EEG theta power during both NREM and REM sleep, and EEG beta power during REM, features suggestive of increased synaptic-metabolic release.

During the sleep deprivation (awake) condition, the **neuro-glymphatic model** with the optimal predictor ***Physio***_***W***_ accounted for over 70% of np-tau and p-tau variance in amyloid-positive individuals, and between 70% and 90% of Aβ and p-tau variance in amyloid-negative individuals. HRV, PTT, and R_P_ contributed to the predictor ***Physio***_***W***_ in a manner largely consistent with their effects during sleep on glymphatic clearance, although R_P_ showed a substantially smaller contribution. In contrast to the optimal sleep predictor, however, EEG delta, theta and beta power that are associated with increased synaptic-metabolic production during wake, contributed strongly to ***Physio***_***W***_.

When we tested the **neuro-glymphatic model** using the single-index predictor ***Hypno***_***S***_ for the sleep condition, constructed from sleep stage durations, it did not outperform the null model in explaining Aβ40 and Aβ42 levels in amyloid-negative individuals, although it did explain over 64% of the variance in tau level. In amyloid-positive individuals, ***Hypno***_***S***_ explained over 30% of the variance. Increased NREM (N2+N3) sleep duration, associated with enhanced glymphatic clearance, and decreased REM sleep, N1 sleep, and WASO, associated with increased synaptic-metabolic release, contributed to ***Hypno***_***S***_. That sleep stage duration alone showed poorer performance in explaining overnight changes in plasma Aβ and tau levels is not surprising. Measuring variations in the underlying neurophysiology occurring in each sleep stage is likely to be more predictive than simply quantifying time spent in each stage.

While it is widely appreciated that glymphatic clearance is more rapid during sleep than during waking, glymphatic clearance does occur during wake^13,17^. Yet to what extent this clearance is modifiable by sleep-like physiology during waking has remained unknown. Sleep deprivation also increases Aβ and tau release into the ISF^29–31^. The wake prediction model using ***Physio***_***W***_ was notable in that it explained a substantial amount of the overnight variance in plasma Aβ and tau levels, with contributions from both increased glymphatic clearance and increased release. The optimal single predictor, ***Physio***_***W***_, showed that changes in cerebrovascular compliance, sympathetic/noradrenergic tone, and parenchymal resistance contributed to increased glymphatic clearance, similar to their effects during sleep in ***Physio***_***S***_. In contrast, wake EEG theta and delta power were associated with increased synaptic-metabolic release. While increased glymphatic clearance during wakefulness contributed to morning Aβ and tau levels, simulated changes in the **neuro-glymphatic model’s** single-index predictor ***Physio***_***W***_ produced predicted shifts in plasma AD biomarker levels that were consistent with release being the primary driver. This interpretation aligns with the multicompartment model, which showed similar results when comparable magnitude changes were made to the cell-to-ISF release rate constant.

We were further able to show from these data that a simulated change in the **neuro-glymphatic model *Physio***_***S***_ or ***Hypno***_***S***_ single-index predictors led to predicted changes in Aβ and tau clearance to plasma that replicated the predictions made by the multicompartment model when similar magnitude changes were made to the CSF-ISF rate constants. Considering that the multicompartment model was derived from external published data^27,28^, this finding independently corroborates the findings from our experimental data and statistical model. It is also noteworthy that the interaction between the single-index predictor and evening plasma concentration in the *experimental* **neuro-glymphatic model** mirrored the interaction between the CSF-ISF and cellular release rate constants and evening plasma concentration in the *multicompartment* **neuro-glymphatic model**. This suggests an association between these rate constants and the neurophysiological or hypnogram measures identified by the single-index predictors. Together these results affirm the **neuro-glymphatic model**, that increased synaptic-metabolic release and glymphatic clearance during sleep supports overnight changes in plasma Aβ and tau levels. These findings further support the predominance of sleep-active glymphatic clearance of Aβ and tau from the brain during sleep, and highlight a key role for synaptic-metabolic production of Aβ and tau during sleep-deprivation.

In addition to supporting the role for glymphatic transport in the clearance of Aβ and tau during sleep, the present results provide additional mechanistic insights into how these processes unfold in the human brain. Consistent with prior studies in rodents and humans, EEG delta power^13,17,22,23^ during NREM sleep and increased cerebrovascular compliance^12,25,40^ contributed to increasing clearance of Aβ and tau to the plasma in the overnight period, while higher EEG theta and delta power during REM sleep contributed to increasing synaptic-metabolic release^29–31^. These contributions are in marked contrast to the dominant effect that R_P_ had within these models, with lower overnight R_P_ contributing to greater morning plasma Aβ and tau levels, which may be because R_P_ mediates the downstream effect of these variables. Conceptually, the synchronized low-frequency neural activity reflected in delta band power and increases in cerebrovascular compliance may serve as a key driving force for the process of glymphatic clearance, while astroglial-mediated changes in extracellular volume fraction reflected in the R_P_ parameter^13,17^ may regulate the effects of this driving force on fluid and solute transport within the brain parenchyma. If true, then R_P_ may represent a promising therapeutic target to increase glymphatic clearance of Aβ and tau.

It is important to point out the distinction between the changes in plasma AD biomarker levels observed through a single overnight period in the present study and changes in plasma AD biomarker levels that reflect steady-state alterations in AD-related pathology. For example, over the timescale of years, declining plasma Aβ42 levels and increasing p-tau levels occur with increasing Aβ plaque and tau pathological burden in AD. Within the setting of the present study, overnight changes in plasma AD biomarker levels reflect the dynamics of both the release of Aβ and tau species into the brain interstitium, and their subsequent clearance to the CSF and the plasma in the overnight period. These dynamics are superimposed upon the existing steady-state Aβ42 and p-tau levels of each participant that were analyzed separately for amyloid-positive and amyloid-negative individuals. Within these two groups, we do not expect individual overnight differences in Cell-ISF release and CSF-ISF exchange to be affected by baseline Aβ42 and p-tau levels. The converse question – whether impaired overnight Aβ and tau clearance leads to increased AD pathological burden and long-term shifts in steady-state plasma AD biomarker levels – is a compelling hypothesis that requires a further investigation.

Our findings showed that elements of sleep-active physiology, in particular, brain parenchymal resistance, enhanced the clearance of Aβ and tau into plasma during overnight sleep in humans. In contrast, during sleep deprivation, increased synaptic–metabolic release elevated plasma AD biomarker levels overnight. Our **null model** and **neuro-glymphatic model** were estimated to predict morning AD biomarker levels from evening AD biomarker levels which is statistically superior to estimating those models on morning-to-evening differences or ratios of AD biomarker levels^63^. We further showed that while synaptic-metabolic release was dominant in the awake state, sleep-related physiology also contributed to increasing Aβ and tau clearance to the plasma.

## SUPPLEMENTAL METHODS AND RESULTS

### Derivation of compartmental pharmacokinetic model

The movement of solute across the compartments in **Figure 1A** can be modeled using first-order kinetics with the published rate constants shown in **Table 1**. This leads to the following linear system of first-order differential equations where f is the cellular production of APP or tau that was assumed to be constitutive, time-independent, across sleep/wake cycles^29,30^:

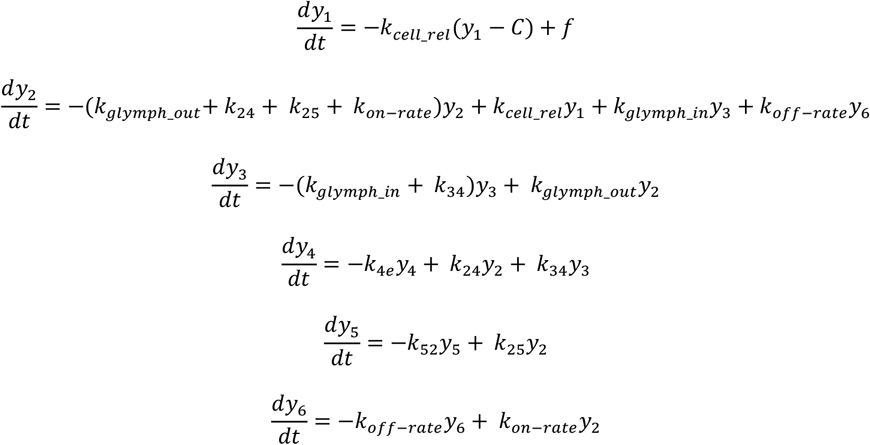

and in matrix notation,

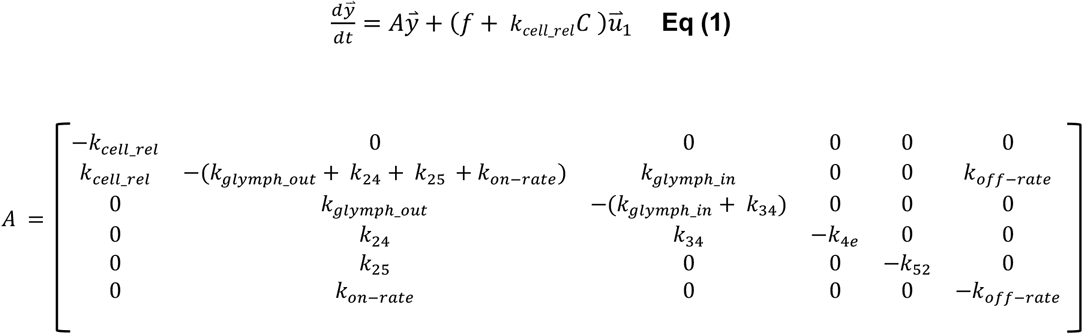

where 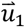 is the unit vector (1,0,0,0,0,0).

**Equation 1** was solved numerically using the *mrgSolve* in *R*. Peptides and proteins released from the Cell compartment to enter the ISF will be cleared from the ISF to the various compartments in a time-dependent manner modeled by 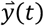. The **null model** assumes, in the absence of sleep-active glymphatic exchange between the ISF and CSF, that the rate constants *k*_*glymph*_*out*_ and *k*_*glymph*_*in*_ are not time dependent. This model further assumes that the rate of release *k*_*cell*_*rel*_ of Aβ and tau from the cell to the ISF compartment during sleep was constant at 30% lower than during wake, and during sleep deprivation was 10% lower than during wake due to circadian regulation.

In contrast, the **neuro-glymphatic model** relaxes the constraint of time-independent rate constants *k*_*glymph*_*out*_ and *k*_*glymph*_*in*_, which are now allowed to increase during sleep, reflecting the increased CSF solute influx and interstitial solute efflux observed in the rodent and human brain^13,16,17^. Furthermore, the Cell release rate *k*_*cell*_*rel*_ of Aβ and tau into the ISF is allowed to vary during sleep and sleep-deprivation reflecting individual changes in synaptic-metabolic production^29–31^.

We then solved the simulation model for 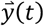 under the **null model** condition and then under the **neuro-glymphatic model** under two scenarios. The first scenario increased the rate constants *k*_*glymph*_*out*_ and *k*_*glymph*_*in*_, and separately increased the Cell release rate constant *k*_*cell*_*rel*_ to define whether morning plasma Aβ and tau levels increase with more rapid overnight glymphatic CSF-ISF exchange or with greater overnight synaptic-metabolic activity.

Of final interest is the solution of a linear approximation of **Equation 1** for the morning compartment concentrations 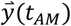 as a function of the evening compartment concentrations 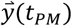. With overnight rate constants *k*_*glymph*_*out*_, *k*_*glymph*_*in*_ and *k*_*cell*_*rel*_ in the **neuro-glymphatic model** held at either increased or decreased level relative to the **null model**, and letting Δ*t* = *t*_*AM*_ - *t*_*PM*_ represent the fixed time between the two blood samples, we apply the fundamental matrix and variation of parameters to solve **Equation 1**, yielding

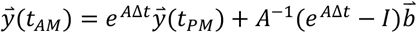

where 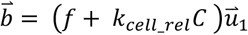. The **neuro-glymphatic model** allows for *k*_*glymph*_*out*_, *k*_*glymph*_*in*_ and *k*_*cell*_*rel*_ in *A* to vary around their fixed values in the **null model**. We are interested in the effect that varying these rate constants has on 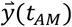. Letting *∂A* denote a matrix of small variations of *k*_*glymph*_*out*_, *k*_*glymph*_*in*_ and *k*_*cell*_*rel*_

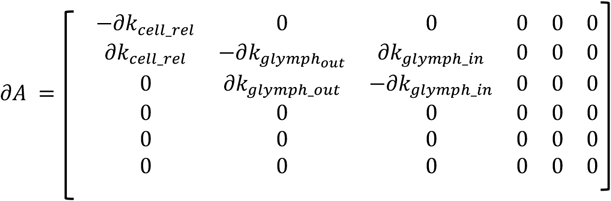

we solve for the first-order approximation of

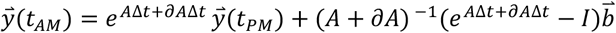

Using the Frechet derivative, a first order approximation gives

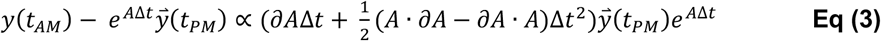

We observe that 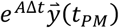 is the multicompartment **null model** prediction of 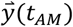 and the right-hand side of **Eq (3)** gives the first-order contribution to 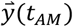 of the **neuro-glymphatic model** solution of the multi-compartment model that arises from varying the rate constants *k*_*glymph*_*out*_, *k*_*glymph*_*in*_ and *k*_*cell*_*rel*_. We note that this effect is linear in *k*_*glymph*_*out*_, *k*_*glymph*_*in*_ and *k*_*cell*_*rel*_, capturing the interaction between evening plasma biomarkers levels 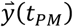 and both glymphatic CSF-ISF exchange and synaptic-metabolic cellular release in the **neuro-glymphatic model** versus the **null model**. In summary, in the **null model**, morning plasma levels are linearly dependent on evening plasma levels only, whereas the **neuro-glymphatic model** has a contribution from the interaction term between the evening plasma levels and the variable rate constants *k*_*glymph*_*out*_, *k*_*glymph*_*in*_ and *k*_*cell*_*rel*_.

### Plasma AD biomarker assessment

The *APOE* genotyping, Aβ and tau plasma biomarkers were analyzed using mass spectrometry by C_2_N Diagnostics^64^. The sample collection procedure was provided by C_2_N Diagnostics. Venipuncture and blood draw from the antecubital fossa were performed using a 22-gauge butterfly needle to minimize red blood cell hemolysis. A total of 10 ml of blood was drawn into a K_2_ EDTA Vacutainer. The blood was centrifuged for 15 min using a swinging bucket rotor at 500-700 x g with the brake on. Immediately after centrifugation, four 1.0 mL plasma samples were aliquoted into four Sarstedt 2.0 ml Micro Tubes without disrupting the plasma/cell interface when transferring plasma. A calibrated air-displacement hand-held pipette with a polypropylene pipette tip was used. After aliquoting plasma into the Sarstedt Micro Tubes, the tubes were immediately capped and frozen at −40°C. When the tubes were ready to be shipped to C_2_N Diagnostics, they were packed into a plastic zip-lock bag with plenty of dry ice, placed in an absorbent towel **and** cryobox, and express couriered to C_2_N Diagnostics priority overnight.

### Validation of PTT measures of cerebrovascular function

The cerebrovascular pulse transit time (PTT) was computed using data acquired from the impedance plethysmogram (IPG) and the left-ear photoplethysmogram (PPG) sensors of the investigational device^17^. A digital filter was applied to the signal in both forward and backward directions using Python SciPy’s filtfilt function, resulting in zero phase distortion. The signal was then bandpass-filtered between 0.5 Hz and 10 Hz using a finite impulse response (FIR) filter with a Hann window (firwin in Python SciPy).

The signal troughs of the IPG measured intracranial cerebrovascular pulsations, while the PPG troughs captured peripheral vascular pulsations. The time difference between the IPG and PPG troughs was calculated as the beat-to-beat cerebrovascular PTT. These intervals were averaged over 30-second EEG epochs.

For the sleep condition, cerebrovascular PTT was expressed as the overnight mean of epoch PTTs normalized to the average PTT of the first three sleep epochs. For the sleep deprivation/wake condition, cerebrovascular PTT was similarly averaged and normalized using the first three wake epochs. Peripheral vascular PTT was computed analogously, using the time difference between the left-ear and right-ear PPG signals^65^. Cerebrovascular PTT during NREM sleep was calculated exclusively over N2 and N3 sleep epochs.

Cerebral blood volume (CBV) was measured the following morning after completion of either the overnight sleep or sleep-deprivation period. A contrast-enhanced, steady-state MRI technique was employed, with normalization to the superior sagittal sinus to obtain absolute CBV values^66^. CBV was measured in eight regions of interest (ROIs): the gray and white matter of the frontal, parietal, temporal, and occipital lobes.

#### Hemodynamic Relationships and Physiological Interpretation

The relationship between mean arterial pressure (MAP) and peripheral PTT is well-established and inversely proportional:

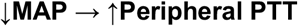

The relationship between MAP and cerebrovascular PTT is similar but more complex due to cerebral vascular autoregulation, which maintains constant cerebral blood flow (CBF) over a wide range of cerebral perfusion pressures (CPP). CPP is defined as:

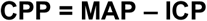

where ICP is intracranial pressure.

Due to autoregulation:

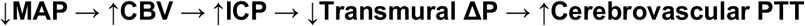

An increase in CBV raises ICP. When this is combined with a decreased MAP, the resulting drop in transmural pressure leads to increased cerebrovascular PTT.

In contrast, when MAP increases:

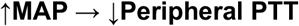

In the peripheral vasculature, higher MAP increases transmural pressure, reducing PTT. In the cerebrovascular system, increased MAP leads to:

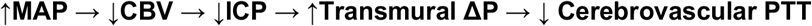

Here, cerebral autoregulation reduces CBV (and ICP) in response to higher MAP, increasing transmural pressure and reducing cerebrovascular PTT.

These mechanisms assume constant brain metabolic demand, under which CBV and cerebrovascular PTT are positively correlated. However, when brain metabolic demand decreases, such as during NREM sleep (assuming MAP remains unchanged):

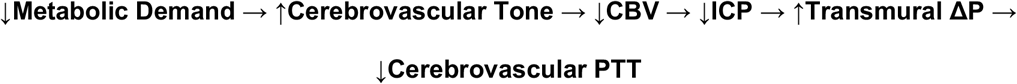

In this case, decreased metabolic demand reduces CBF via increased vascular tone, leading to reduced CBV and ICP, which in turn increases transmural pressure and reduces cerebrovascular PTT.

Thus, we conclude that cerebral autoregulation maintains a positive association between cerebral blood volume (CBV) and cerebrovascular pulse transit time (PTT) under both fluctuations in mean arterial pressure (MAP) and changes in brain metabolic demand. However, it is important to note that MAP and metabolic demand can vary independently or in opposing directions.

**Supplemental Table 2** presents the overnight sleep and wake peripheral PTT and cerebrovascular PTT in amyloid-positive and amyloid-negative participants. While the sleep-to-wake differences are not statistically significant, likely due to the small sample size, the observed trends offer valuable insights.

Among amyloid-negative participants, peripheral PTT increased during sleep compared to wakefulness. This pattern aligns with a decrease in MAP during sleep, which, assuming metabolic demand remains constant, would increase CBV and, consequently, increase cerebrovascular PTT. However, sleep is also characterized by reduced brain metabolic demand, which tends to decrease CBV and lower cerebrovascular PTT. These opposing physiological effects appear to offset each other, resulting in minimal net change in cerebrovascular PTT between sleep and wake in this group.

In contrast, amyloid-positive participants showed a decrease in peripheral PTT during sleep relative to wakefulness. This trend suggests an increase in MAP during sleep, which would reduce CBV. Combined with the reduction in metabolic demand during sleep, which also lowers CBV, the additive effect would be a decrease in cerebrovascular PTT, a finding that is consistent with the values shown in **Supplemental Table 2**.

To test the hypothesis that overnight cerebrovascular PTT predicts morning CBV, we employed a multivariate linear mixed-effects model. This model included eight dependent CBV variables, one for each region of interest (ROI), and a single independent predictor: cerebrovascular PTT.

Potential confounding variables were included in the model: age, sex, APOE E4 carrier status, and circadian alignment, as previously described. All participants were drawn from the Benchmarking Study; participants in the Replication Study were excluded from this analysis, as they did not undergo contrast-enhanced MRI and therefore had no CBV measurements available.

The regression coefficients for cerebrovascular PTT in the multivariate linear mixed model were positive and statistically significant across all eight ROIs for amyloid-negative participants, both following overnight sleep and overnight sleep deprivation (**Supplemental Tables 3 and 4**). These findings are consistent with the proposed positive physiological relationship between CBV and cerebrovascular PTT.

In amyloid-positive participants, the cerebrovascular PTT coefficients were also positive and significant across all eight ROIs following overnight sleep deprivation, and positive and significant for the four gray matter ROIs following overnight sleep (**Supplemental Tables 3 and 4**).

Model performance was evaluated by comparing the prediction model, which included cerebrovascular PTT, against a null model that excluded cerebrovascular PTT. The likelihood ratio test (LRT) indicated that the prediction model provided a significantly better fit across all four comparisons: amyloid-positive vs. amyloid-negative and sleep vs. sleep deprivation (**Supplemental Table 5**).

Using the marginal variances for each model that reflect the total population-level variance of between-subject variation and within-subject variation, the prediction model explained between 63.8% and 85.4% of the variance attributed to the **null model** across the eight regions of interest (ROIs) in each of the four comparisons^67^. Similarly high percentages of variance were explained when analyzed by individual ROI (**Supplemental Table 6**).

#### Construction of Single Predictors

The objective of the single-predictor approach, using single-index regression^34^, was to identify a single linear combination of predictors that would allow inferences about the relative contributions of glymphatic clearance and synaptic-metabolic release variables in explaining plasma biomarker levels within the **neuro-glymphatic model**. Given the inferential nature of the hypotheses, all variables were retained, and models were selected based on maximum log-likelihood. The construction proceeded through two steps: **1)** identifying a maximal non-collinear subset of the predictor regressors from the appropriate group of regressors; **2)** running an optimization algorithm to find the weights for the linear combination that minimized the log-likelihood of the **neuro-glymphatic model** on the data^68^. Optimization was performed using the R optim function with Nelder-Mead optimization. Each optimization was run with multiple initial parameter values and the best result selected. Once the best single predictor subset of regressors and weights were determined for ***Physio***_***S***_, ***Physio***_***W***_ and ***Hypno***_***S***_, Wald’s p values and 95% CI were estimated on each of the weights using 1,000 bootstrap re-samples. The R package *lmeresampler* for bootstrap routines for nested linear mixed effects models was used to resample the data.

### Multivariate model representing the null model and neuro-glymphatic models

The R lme4 package was used for the linear mixed models. The multivariate linear mixed model was modeled in lme4^69^.

The **null model** represented the null hypothesis that efflux of Aβ and tau from the ISF to the CSF is not dependent on glymphatic exchange, that is, *k*_*glymph*_*out*_ and *k*_*glymph*_*in*_ are time-invariant. Furthermore, in this model efflux of Aβ and tau into the ISF from synaptic-metabolic activity, *k*_*cell*_*rel*_, decreases with sleep but remains time-invariant during sleep. In this model, morning plasma levels of Aβ and tau species are assumed not to be influenced by either of the two groups (***Physio, Hypno***) of sleep-related predictors. Letting ***Conc***_***AM***_ and ***Conc***_***PM***_ denote the morning and evening plasma levels of the five Aβ and tau biomarkers Aβ_40_, Aβ_42_, np-tau181, np-tau217 and p-tau181, respectively, the **null model** is the multivariate linear mixed model that regresses ***Conc***_***AM***_ on ***Comc***_***PM***_ with covariates of amyloid status, age, APOE-ε4 status, sex, circadian rhythm and study site, with participant ID as random intercept. This multivariate model was solved using R library lme4 after transforming the ***Conc***_***AM***_ and ***Conc***_***PM***_ into one-dimensional vectors and adding a variable *biomarker* to index these observations in the original data^69^. The categorical *biomarker* variable was included in the model as an interaction term with the transformed ***Conc***_***PM***_ and as a vector of random slopes. Specifically, random effects were specified as (biomarker - 1 I id) in **lme4** syntax, yielding a full (unstructured) covariance matrix among the five biomarker-specific slopes. This formulation allowed correlations between biomarker variables to be estimated via their shared subject-level covariance structure, effectively modeling cross-biomarker dependencies within participants. ***Conc***_***AM***_ and ***Conc***_***PM***_ were rescaled to improve numerical stability of the linear mixed model estimation. Specifically, values for Aβ_40_ were divided by 10 so that regression coefficients reflect the effect of a 10-unit increase in the original scale.

The **neuro-glymphatic model** represented the alternative hypothesis that clearance of Aβ and tau species from the ISF to plasma is dependent on (i) sleep-active glymphatic exchange, with *k*_*glymph*_*out*_ and *k*_*glymph*_*in*_ increasing overnight in the sleep condition; and (ii) variation in synaptic-metabolic Aβ and tau species release into the ISF through overnight changes in *k*_*cell*_*rel*_. Three glymphatic models were tested, two for the sleep condition with the first, ***Physio***_***S***,_ using features of parenchymal resistance, sleep neurophysiology, central noradrenergic tone, and cerebrovascular function while the second, ***Hypno***_***S***_, reflects only hypnographic sleep stage duration. For the sleep deprivation/wake condition, ***Physio***_***W***_, used the same features as ***Physio***_***S***_. The predictors ***Physio***_***S***_ and ***Physio***_***W***_ denote single predictors constructed from the linear combination of R_p_, HRV, PTT, and EEG power bands during the sleep and sleep deprivation (wake) conditions, respectively. The predictor ***Hypno***_***S***_ denotes the single predictor constructed from the linear combination of REM, NREM, N1 and WASO hypnogram stage duration. The two sleep **neuro-glymphatic models** added to the **null model** the predictors ***Physio***_***S***_ and ***Hypno***_***S***_ and their respective interaction terms with evening levels of plasma Aβ and tau ***Physio***_***S***_ * ***Conc***_***PM***_ and ***Hypno***_***S***_ ***\* Conc***_***PM***_. The sleep deprivation/wake **neuro-glymphatic model** added to the **null model** the predictor ***Physio***_***W***_ and the interaction term ***Physio***_***W***_ * ***Conc***_***PM***_. The R *Anova* function in the car package was used for analysis of deviance using a Type III Wald Chi-square test for the interaction effects.

The likelihood ratio test (LRT) was used for significance testing of the **neuro-glymphatic model** versus the **null model**. Of particular interest was the percent of the residual variation in the **null model** that was explained by the **glymphatic model**; that is, the additional variation explained by including one of the single predictors ***Physio***_***S***_, ***Physio***_***W***_ or ***Hypno***_***S***_ in the **neuro-glymphatic model** under the normal sleep or sleep deprivation/wake conditions. The residual variation of each model was computed from the sum of the square difference between the observed morning plasma AD biomarker levels and the fitted morning plasma levels for Aβ_40_, Aβ_42_, np-tau181, np-tau217 and p-tau181 individually using maximum likelihood and conditional on the random effects. Letting *RSS*_*O*_ and *RSS* denote these two quantities for the **null model** and a **glymphatic model**, 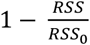 gives the R^2^ of the multivariate linear mixed models^67^. Confidence intervals for these R^2^ estimates were computed from quantiles on 1,000 conditional parametric bootstrap resamples.

### Sensitivity analysis to define individual predictor contributions

We quantified the contribution of each individual predictor to glymphatic clearance or synaptic-metabolic release during sleep by simulating changes in the magnitude of ***Physio***_***S***_. A sensitivity analysis of the individual predictors within ***Physio***_***S***_ was conducted using the **neuro-glymphatic model** (**Table 5**). Morning plasma concentrations of AD biomarkers ***Conc***_***AM***_ were simulated following a 5% increase in the magnitude of each predictor (**Supplemental Table 8**). By multiplying the effect of each predictor by its respective weight in the linear single-predictor ***Physio***_***S***_, we determined that a negative value indicated that a **decrease** in predictor magnitude led to an **increase** in ***Conc***_***AM***_, whereas a positive value indicated that an **increase** in predictor magnitude resulted in a **decrease** in ***Conc***_***AM***_ (**Supplemental Table 8**). Notably, a decrease in ***Physio***_***S***_ led to all individual predictors increasing morning plasma concentrations of AD biomarkers.

To define if this increase in morning plasma AD biomarker levels was attributable to enhanced glymphatic clearance or from increased synaptic-metabolic release, we referenced the numerical sign (+/−) of the individual predictor weights in ***Physio***_***s***_, with (i) a positive sign (+) reflecting that a *decrease* in that predictor contributes to the increase in morning plasma AD biomarker concentrations, and (ii) a negative sign (−) reflecting that an *increase* in that predictor contributes to the increase in morning plasma AD biomarker concentrations. For each of these individual predictors, their presumed effect upon glymphatic clearance and synaptic-metabolic release of Aβ and tau can be defined based on prior literature ^**2,13,16,17,22,23,25,29–31,40,41,70-73**^, as outlined in **Supplemental Table 9**. By combining the numerical signs of the individual predictor weights and the individual predictor’s presumed effects on glymphatic clearance and synaptic-metabolic release of Aβ and tau, the likely mechanism linking the individual predictor to the increase in morning plasma AD biomarker levels could be inferred (**Supplemental Table 8**, right column).

As an example, **Supplemental Table 8** shows that the effect of PTT is to increase ***Conc***_***AM***_ when ***Physio***_***S***_ is decreased, an effect that could be attributable to an increase in brain-to-blood clearance, or increased synaptic-metabolic release of Aβ and tau. Because the numerical sign of the PTT weight estimate is negative (−0.034 for amyloid-positive individuals), its magnitude increases when ***Physio***_***S***_ is decreased. Combining these two results, the effect of an *increase* in PTT is to *increase* ***Conc***_***AM***_. Based on the role that vasomotor oscillations play in perivascular transport and glymphatic clearance^12,13,25,40^, it is presumed that this effect of PTT on ***Conc***_***AM***_ occurs through enhanced clearance by increasing perivascular CSF transport. Using a similar approach, we conclude that sleep-related individual predictors that are associated with increased glymphatic function, such as NREM EEG delta power^13,17,22,23^, cerebrovascular compliance measured by PTT^12,13,25,40^, and R_P_^13,17^ were each associated with increased overnight clearance of Aβ and tau into the plasma. In contrast, NREM EEG theta and REM EEG theta and beta spectral power appear to be linked to elevated production^29–31,73^.

The compartmental pharmacokinetic model predicts that increases in both sleep-active glymphatic function and synaptic-metabolic activity contribute to increased morning plasma AD biomarker levels (**Figure 1A-B**). Consistent with this prediction, as shown in **Supplemental Table 8**, at the mean evening plasma biomarker values, a decrease in overnight ***Physio***_***S***_ corresponded to higher morning plasma AD biomarker levels across all individual predictors. Because each neurophysiologic predictor in ***Physio***_***S***_ was normalized to its value at sleep onset^17^, its weight in the linear combination defining ***Physio***_***S***_ reflects its relative contribution to its primary mechanism driving morning biomarker increases. As evident from **Table 4** and **Supplemental Table 8**, parenchymal resistance R_P_, which is a key determinant of sleep-active glymphatic clearance^13,17^, was the dominant contributor to ***Physio***_***S***_, while other neurophysiological predictors (PTT, HRV, EEG power bands) had comparatively minor effects on either glymphatic clearance or synaptic-metabolic release.

---

**Supplemental Table 1.** Effect of changes in glymphatic clearance or Aβ and tau release on plasma Aβ42/Aβ40 and p-tau/np-tau ratios.

| $k_{on-rate}$ | $k_{off-rate}$ | Change in<br>$k_{glymph\_in}$ and $k_{glymph\_out}$ | Change in<br>$k_{cell\_rel}$ | Increase in Aβ42/ Aβ40 | Increase in p-tau/np-tau |
| --- | --- | --- | --- | --- | --- |
| 0 | - | Any | Any | 0% | 0% |
| 7.2 hr<sup>-1</sup> | 0.32 hr<sup>-1</sup> | 2.0x | - | 4.7% | 4.9% |
| 7.2 hr<sup>-1</sup> | 0.32 hr<sup>-1</sup> | - | 0.5x | 7.4% | 4.4% |
| 1.0 hr<sup>-1</sup> | 0.1 hr<sup>-1</sup> | 2.0x | - | 3.4% | 3.5% |
| 1.0 hr<sup>-1</sup> | 0.1 hr<sup>-1</sup> | - | 0.5x | 5.1% | 2.5% |
| 1.0 hr<sup>-1</sup> | 1.0 hr<sup>-1</sup> | 2.0x | - | 2.4% | 2.6% |
| 1.0 hr<sup>-1</sup> | 1.0 hr<sup>-1</sup> | - | 0.5x | 4.0% | 2.5% |
| 0.1 hr<sup>-1</sup> | 1.0 hr<sup>-1</sup> | 2.0x | - | 0.4% | 0.4% |
| 0.1 hr<sup>-1</sup> | 1.0 hr<sup>-1</sup> | - | 0.5x | 0.7% | 0.5% |
| 7.2 hr<sup>-1</sup> | 0.32 hr<sup>-1</sup> | 0.5x | - | -2.5% | -2.4% |
| 7.2 hr<sup>-1</sup> | 0.32 hr<sup>-1</sup> | - | 1.5x* | -4.9% | -3.1% |
| 1.0 hr<sup>-1</sup> | 0.1 hr<sup>-1</sup> | 0.5x | - | -1.7% | -1.6% |
| 1.0 hr<sup>-1</sup> | 0.1 hr<sup>-1</sup> | - | 1.5x | -3.4% | -1.9% |
| 1.0 hr<sup>-1</sup> | 1.0 hr<sup>-1</sup> | 0.5x | - | -1.4% | -1.3% |
| 1.0 hr<sup>-1</sup> | 1.0 hr<sup>-1</sup> | - | 1.5x | -2.6% | -1.8% |
| 0.1 hr<sup>-1</sup> | 1.0 hr<sup>-1</sup> | 0.5x | - | -0.2% | -0.2% |
| 0.1 hr<sup>-1</sup> | 1.0 hr<sup>-1</sup> | - | 1.5x | -0.5% | -0.3% |

\*A 1.5x increase in $k_{cell\_rel}$ during sleep put it at 105% of its value at wake.

**Supplemental Table 2:**
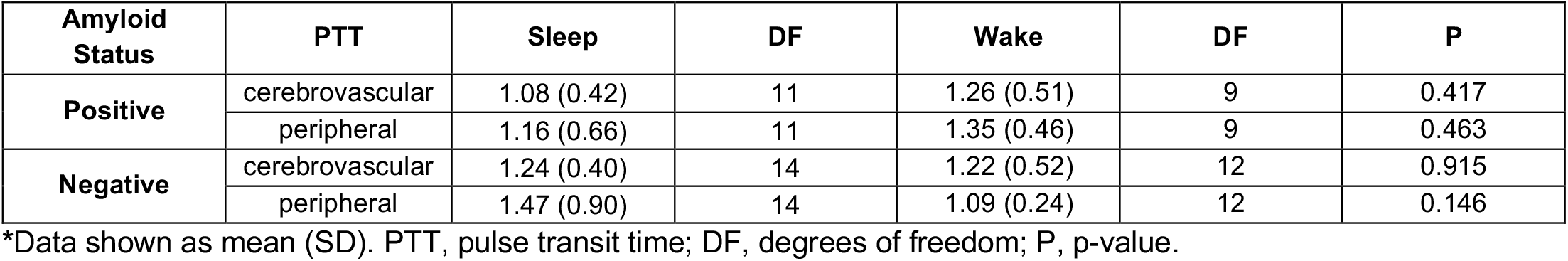
Cerebrovascular and peripheral PTT during sleep and wake visits.

**Supplemental Table 3:** Prediction of morning CBV for gray and white matter ROI following overnight sleep with NREM cerebrovascular PTT *(prediction model)* and without *(null model)* in amyloid positive and amyloid negative individuals*.

| Model | Predictor Group | Predictor |  | Amyloid Positive |  |  |  | Amyloid Negative |  |  |  |
| --- | --- | --- | --- | --- | --- | --- | --- | --- | --- | --- | --- |
|  |  |  |  | Estimate | DF | CI | P | Estimate | DF | CI | P |
| Null Model | Fixed Effects | Age |  | 0.289 | 46 | 0.287 – 0.292 | **<0.001** | -0.008 | 70 | -0.023 – 0.007 | 0.313 |
|  |  | Sex (male) |  | -0.427 | 46 | -0.438 – -0.417 | **<0.001** | 0.488 | 70 | 0.381 – 0.596 | **<0.001** |
|  |  | APOE-ε4 (positive) |  | 1.501 | 46 | 1.489 – 1.512 | **<0.001** | -0.398 | 70 | -0.519 – -0.278 | **0.047** |
|  |  | Circadian |  | -0.033 | 46 | -0.033 – -0.032 | **<0.001** | 0.000 | 70 | -0.001 – -0.000 | 0.343 |
| | Random Effects | $\sigma^2$ | | 0 | | | | 0.001 | | | |
|  |  | T11 pid.Frontal GM |  | 0.745 |  |  |  | 4.028 |  |  |  |
|  |  | T11 pid.Frontal WM |  | 2.636 |  |  |  | 0.275 |  |  |  |
|  |  | T11 pid.Occipital GM |  | 29.092 |  |  |  | 79.264 |  |  |  |
|  |  | T11 pid.Occipital WM |  | 3.55 |  |  |  | 0.564 |  |  |  |
|  |  | T11 pid.Parietal GM |  | 2.895 |  |  |  | 15.981 |  |  |  |
|  |  | T11 pid.Parietal WM |  | 2.539 |  |  |  | 0.146 |  |  |  |
|  |  | T11 pid.Temporal GM |  | 5.276 |  |  |  | 20.111 |  |  |  |
|  |  | T11 pid.Temporal WM |  | 2.415 |  |  |  | 0.275 |  |  |  |
| Prediction Model | Fixed Effects | Age |  | 0.27 | 38 | 0.269 – 0.271 | **<0.001** | -0.021 | 62 | -0.025 – -0.016 | **<0.001** |
|  |  | Sex (male) |  | -0.304 | 38 | -0.308 – -0.3 | **<0.001** | 0.218 | 62 | 0.179 – 0.256 | **<0.001** |
|  |  | APOE-ε4 (positive) |  | 1.345 | 38 | 1.341 – 1.35 | **<0.001** | -0.385 | 62 | -0.425 – -0.346 | **<0.001** |
|  |  | Circadian |  | -0.032 | 38 | -0.032 – -0.032 | **<0.001** | -0.001 | 62 | -0.002 – -0.001 | **<0.001** |
|  |  | Predictor Effects | 0.157 | 0.970 | 38 | -0.25 – 0.564 | 0.463 | 2.046 | 62 | 1.556 – 2.536 | **<0.001** |
|  |  |  | -1.328 | -0.516 | 38 | -1.792 – -0.864 | **<0.001** | 0.489 | 62 | 0.296 – 0.682 | **<0.001** |
|  |  |  | 5.151 | 5.964 | 38 | 3.513 – 6.789 | **<0.001** | 6.991 | 62 | 5.311 – 8.671 | **<0.001** |
|  |  |  | -1.589 | -0.776 | 38 | -2.112 – -1.066 | **<0.001** | 0.383 | 62 | 0.12 – 0.646 | **0.012** |
|  |  |  | 1.612 | 2.424 | 38 | 1.058 – 2.166 | **<0.001** | 3.489 | 62 | 2.671 – 4.308 | **<0.001** |
|  |  |  | -1.382 | -0.570 | 38 | -1.795 – -0.97 | **<0.001** | 0.526 | 62 | 0.378 – 0.674 | **<0.001** |
|  |  |  | 2.128 | 2.941 | 38 | 1.363 – 2.894 | **<0.001** | 3.849 | 62 | 2.953 – 4.745 | **<0.001** |
|  |  |  | -1.201 | -0.388 | 38 | -1.684 – -0.718 | **<0.001** | 0.634 | 62 | 0.366 – 0.902 | **<0.001** |
| | Random Effects | $\sigma^2$ | | 0 | | | | 0.000 | | | |
|  |  | T11 pid.Frontal GM |  | 0.655 |  |  |  | 1.444 |  |  |  |
|  |  | T11 pid.Frontal WM |  | 0.85 |  |  |  | 0.215 |  |  |  |
|  |  | T11 pid.Occipital GM |  | 10.589 |  |  |  | 17.078 |  |  |  |
|  |  | T11 pid.Occipital WM |  | 1.079 |  |  |  | 0.408 |  |  |  |
|  |  | T11 pid.Parietal GM |  | 1.211 |  |  |  | 4.048 |  |  |  |
|  |  | T11 pid.Parietal WM |  | 0.671 |  |  |  | 0.122 |  |  |  |
|  |  | T11 pid.Temporal GM |  | 2.311 |  |  |  | 4.851 |  |  |  |
|  |  | T11 pid.Temporal WM |  | 0.919 |  |  |  | 0.425 |  |  |  |

\*CBV, cerebral blood volume; ROI, region of interest; GM, gray matter; WM, white matter; DF, degrees of freedom; CI, 95% confidence interval; P, p-value; PTT, cerebrovascular pulse-transit time measured during NREM sleep; $\sigma^2$ , residual variance; $\tau_{11}$ , random slope variance; **Bolded P values reflect significant associations at P < 0.05**. Shaded cells demark random effects and potential biological confounders.

**Supplemental Table 4:**
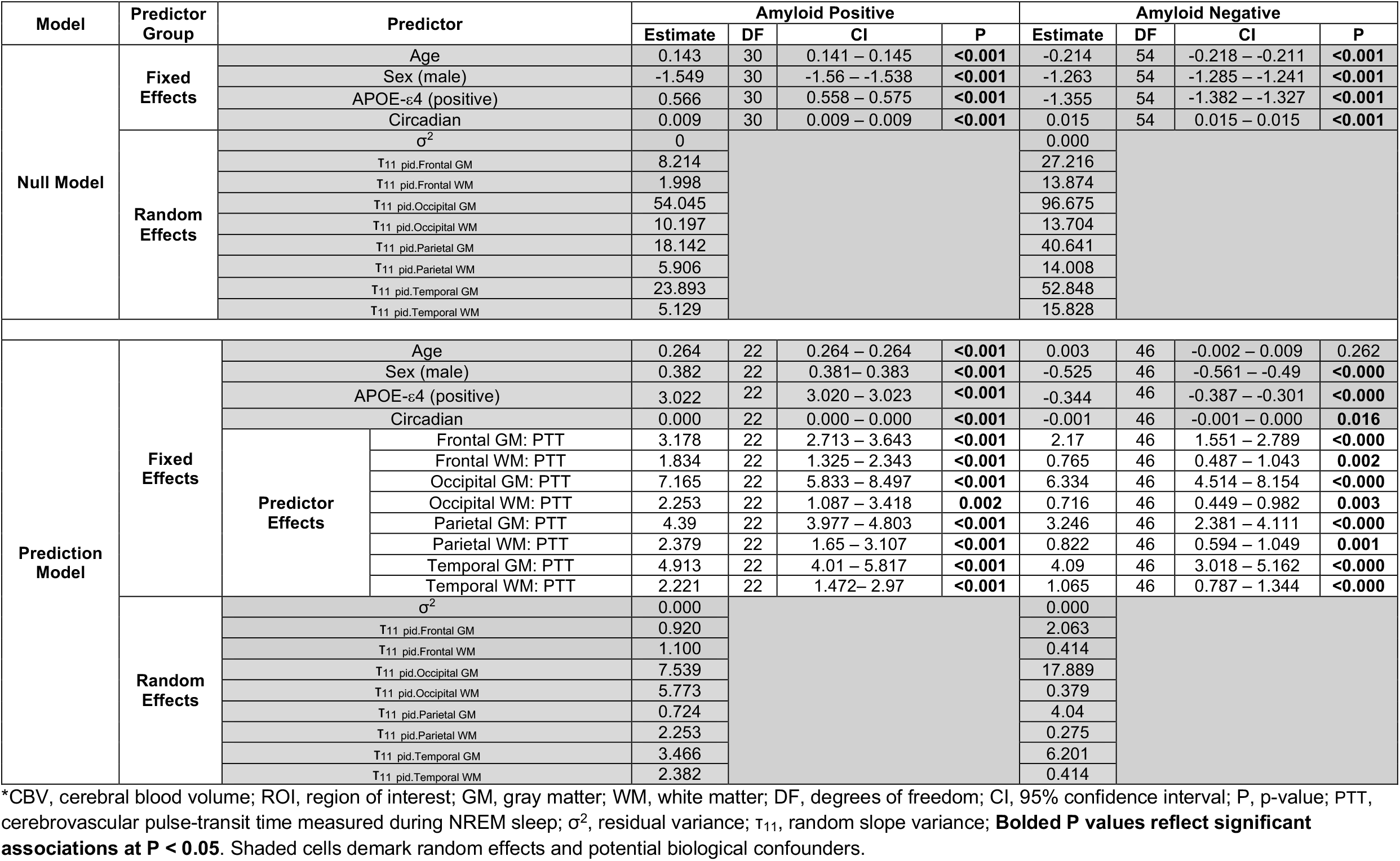
Prediction of morning CBV for gray and white matter ROI following overnight sleep deprivation (wake) with cerebrovascular PTT *(prediction model)* and without *(null model)* in amyloid positive and amyloid negative individuals*.

**Supplemental Table 5:** Likelihood ratio test of the *prediction model* versus the *null model*\*.

| Condition | Amyloid Status | Model | AIC | BIC | logLik | LRT | DF | P |
| --- | --- | --- | --- | --- | --- | --- | --- | --- |
| Sleep Condition | Positive | Null | 117.9 | 221.9 | -16.9 |  |  |  |
|  |  | Prediction | 81.3 | 205.2 | 9.3 | 52.5 | 8 | **<0.001** |
|  | Negative | Null | 247.9 | 362.1 | -81.9 |  |  |  |
|  |  | Prediction | 202.8 | 338.7 | -51.4 | 61.1 | 8 | **<0.001** |
| Sleep Deprivation/Wake | Positive | Null | 166.7 | 262.4 | -41.4 |  |  |  |
|  |  | Prediction | 113.9 | 227.7 | -7.0 | 68.8 | 8 | **<0.001** |
|  | Negative | Null | 153.3 | 261.0 | -34.6 |  |  |  |
|  |  | Prediction | 149.0 | 277.2 | -24.5 | 20.3 | 8 | **0.009** |

\*AIC, Akaike Information Criteria; BIC, Bayesian Information Criteria; logLik, log likelihood; LRT, log likelihood ratio; DF, degrees of freedom; P, p-value; **Bolded P values reflect significant LRT at P < 0.05.**

**Supplemental Table 6.**
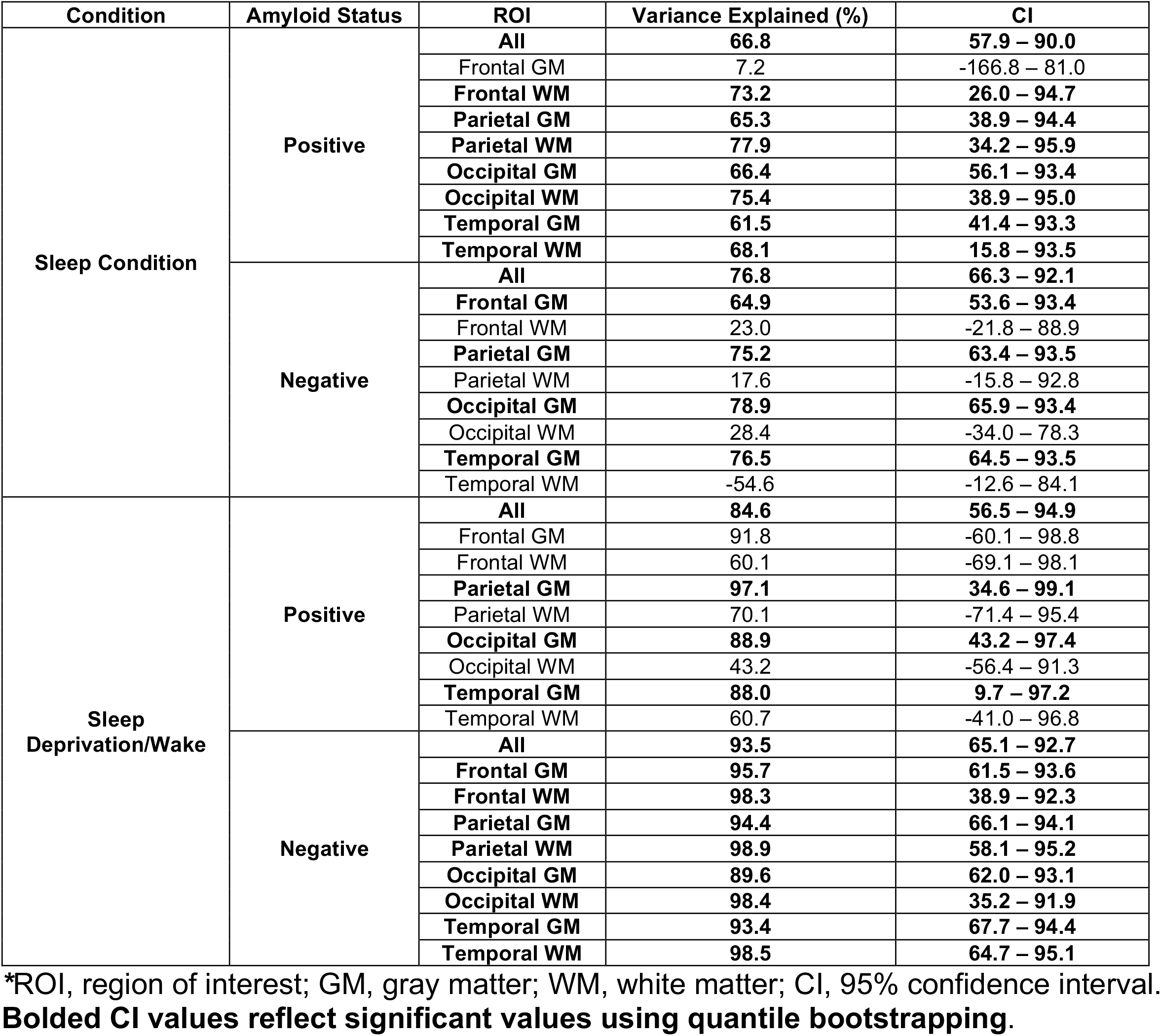
Percent of variance in morning plasma AD biomarker levels explained by the *neuro-glymphatic model* over the *null model*\*.

| Condition | Amyloid Status | ROI | Variance Explained (%) | CI |
| --- | --- | --- | --- | --- |
| Sleep Condition | Positive | All | 66.8 | 57.9 – 90.0 |
|  |  | Frontal GM | 7.2 | -166.8 – 81.0 |
|  |  | Frontal WM | 73.2 | 26.0 – 94.7 |
|  |  | Parietal GM | 65.3 | 38.9 – 94.4 |
|  |  | Parietal WM | 77.9 | 34.2 – 95.9 |
|  |  | Occipital GM | 66.4 | 56.1 – 93.4 |
|  |  | Occipital WM | 75.4 | 38.9 – 95.0 |
|  |  | Temporal GM | 61.5 | 41.4 – 93.3 |
|  |  | Temporal WM | 68.1 | 15.8 – 93.5 |
|  | Negative | All | 76.8 | 66.3 – 92.1 |
|  |  | Frontal GM | 64.9 | 53.6 – 93.4 |
|  |  | Frontal WM | 23.0 | -21.8 – 88.9 |
|  |  | Parietal GM | 75.2 | 63.4 – 93.5 |
|  |  | Parietal WM | 17.6 | -15.8 – 92.8 |
|  |  | Occipital GM | 78.9 | 65.9 – 93.4 |
|  |  | Occipital WM | 28.4 | -34.0 – 78.3 |
|  |  | Temporal GM | 76.5 | 64.5 – 93.5 |
|  |  | Temporal WM | -54.6 | -12.6 – 84.1 |
| Sleep Deprivation/Wake | Positive | All | 84.6 | 56.5 – 94.9 |
|  |  | Frontal GM | 91.8 | -60.1 – 98.8 |
|  |  | Frontal WM | 60.1 | -69.1 – 98.1 |
|  |  | Parietal GM | 97.1 | 34.6 – 99.1 |
|  |  | Parietal WM | 70.1 | -71.4 – 95.4 |
|  |  | Occipital GM | 88.9 | 43.2 – 97.4 |
|  |  | Occipital WM | 43.2 | -56.4 – 91.3 |
|  |  | Temporal GM | 88.0 | 9.7 – 97.2 |
|  |  | Temporal WM | 60.7 | -41.0 – 96.8 |
|  | Negative | All | 93.5 | 65.1 – 92.7 |
|  |  | Frontal GM | 95.7 | 61.5 – 93.6 |
|  |  | Frontal WM | 98.3 | 38.9 – 92.3 |
|  |  | Parietal GM | 94.4 | 66.1 – 94.1 |
|  |  | Parietal WM | 98.9 | 58.1 – 95.2 |
|  |  | Occipital GM | 89.6 | 62.0 – 93.1 |
|  |  | Occipital WM | 98.4 | 35.2 – 91.9 |
|  |  | Temporal GM | 93.4 | 67.7 – 94.4 |
|  |  | Temporal WM | 98.5 | 64.7 – 95.1 |

\*ROI, region of interest; GM, gray matter; WM, white matter; CI, 95% confidence interval. **Bolded CI values reflect significant values using quantile bootstrapping.**

**Supplemental Table 7.** Estimate for single predictor coefficient in the *neuro-glymphatic model* in sleep and sleep deprivation evaluated at the mean evening plasma AD biomarker levels*.

| Condition and Predictor | Amyloid Status | Factor | Estimate at Mean Value of $Conc_{PM}$ for Biomarker | P of Main Effect | P of Interaction Effect |
| --- | --- | --- | --- | --- | --- |
| Sleep Condition *Physios* | Positive | $(1+\overline{Conc}_{PM,Ab40})$ *Physios* | -327.245 | **0.035** | **<0.001** |
| | | $(1+\overline{Conc}_{PM,Ab42})$ *Physios* | -297.768 | **0.030** | **<0.001** |
| | | $(1+\overline{Conc}_{PM,Tau})$ *Physios* | -442.340 | **0.015** | **<0.001** |
| | Negative | $(1+\overline{Conc}_{PM,Ab40})$ *Physios* | -5.733 | 0.179 | **0.017** |
| | | $(1+\overline{Conc}_{PM,Ab42})$ *Physios* | -7.652 | **0.001** | **<0.001** |
| | | $(1+\overline{Conc}_{PM,nTau181})$ *Physios* | -7.179 | **<0.001** | **<0.001** |
| | | $(1+\overline{Conc}_{PM,nTau217})$ *Physios* | -6.619 | **<0.001** | **<0.001** |
| | | $(1+\overline{Conc}_{PM,pTau181})$ *Physios* | -3.607 | 0.206 | **<0.001** |
| Sleep Condition *Hypnos* | Positive | $(1+\overline{Conc}_{PM,Ab40})$ *Hypnos* | -0.137 | **0.015** | **0.025** |
| | | $(1+\overline{Conc}_{PM,Ab42})$ *Hypnos* | -0.134 | **0.023** | 0.057 |
| | | $(1+\overline{Conc}_{PM,Tau})$ *Hypnos* | 0.075 | 0.184 | **0.041** |
| | Negative | $(1+\overline{Conc}_{PM,Ab40})$ *Hypnos* | -0.038 | **0.001** | **0.001** |
| | | $(1+\overline{Conc}_{PM,Ab42})$ *Hypnos* | -0.070 | **<0.001** | **<0.001** |
| | | $(1+\overline{Conc}_{PM,nTau181})$ *Hypnos* | -0.058 | **<0.001** | **<0.001** |
| | | $(1+\overline{Conc}_{PM,nTau217})$ *Hypnos* | -0.050 | **<0.001** | **<0.001** |
| | | $(1+\overline{Conc}_{PM,pTau181})$ *Hypnos* | 0.003 | **<0.001** | **<0.001** |
| Sleep Deprivation/Wake *Physio<sub>w</sub>* | Positive | $(1+\overline{Conc}_{PM,Ab40})$ *Physio<sub>w</sub>* | 4.746 | 0.290 | 0.281 |
| | | $(1+\overline{Conc}_{PM,Ab42})$ *Physio<sub>w</sub>* | 5.725 | 0.313 | 0.318 |
| | | $(1+\overline{Conc}_{PM,Tau})$ *Physio<sub>w</sub>* | -51.708 | 0.433 | **<0.001** |
| | Negative | $(1+\overline{Conc}_{PM,Ab40})$ *Physio<sub>w</sub>* | -9.636 | **<0.001** | **0.002** |
| | | $(1+\overline{Conc}_{PM,Ab42})$ *Physio<sub>w</sub>* | -9.682 | **<0.001** | **0.001** |
| | | $(1+\overline{Conc}_{PM,nTau181})$ *Physio<sub>w</sub>* | -4.140 | **<0.001** | **<0.001** |
| | | $(1+\overline{Conc}_{PM,nTau217})$ *Physio<sub>w</sub>* | -4.158 | **<0.001** | **<0.001** |
| | | $(1+\overline{Conc}_{PM,pTau181})$ *Physio<sub>w</sub>* | -3.393 | **<0.001** | **<0.001** |

**$Conc_{PM}$** , evening plasma level for the biomarker; ***Physios*** and ***Physio<sub>w</sub>***, single predictors constructed from the linear combination of the full model predictors during the sleep and sleep deprivation (wake) conditions, respectively, for both amyloid positive and negative participants separately; ***Hypnos***, single predictor constructed from the full model hypnogram features during the sleep condition for both amyloid positive and negative participants separately; **Bolded P values reflect significant LRT at $P < 0.05$ .**

**Supplemental Table 8.** Change in morning plasma levels of AD biomarker levels under sleep and sleep deprivation conditions following a 5% magnitude increase in a predictor*.

| Condition and Predictor | Amyloid Status | Predictors | Estimate | Effect on *Conc<sub>AM</sub>* at Mean Value of *Conc<sub>PM</sub>* Following a 5% Magnitude Increase in Predictor |  |  |  |  | Individual Predictor Effect on *Conc<sub>AM</sub>* when *Physio<sub>s</sub>*, *Hypno<sub>s</sub>*, or *Physio<sub>w</sub>* are Decreased | Increased Clearance or Increased Production Mechanism |
| --- | --- | --- | --- | --- | --- | --- | --- | --- | --- | --- |
|  |  |  |  | *Aβ*40 | *Aβ*42 | *nTau*181 | *nTau*217 | *pTau*181 |  |  |
| Sleep Condition *Physio<sub>s</sub>* | Positive | R<sub>p</sub> | 0.968 | -14.589 | -13.275 | -19.720 |  |  | Increase | Clearance |
|  |  | PTT (NREM) | -0.034 | 0.607 | 0.553 | 0.821 |  |  | Increase | Clearance |
|  |  | HRV (NREM) | -0.068 | 1.581 | 1.439 | 2.138 |  |  | Increase | Clearance |
|  |  | EEG Delta Power (NREM) | 0.086 | -1.915 | -1.743 | -2.589 |  |  | Increase | Production |
|  |  | EEG Theta Power (NREM) | 0.162 | -2.585 | -2.352 | -3.494 |  |  | Increase | Production |
|  |  | EEG Theta Power (REM) | -0.146 | 2.467 | 2.245 | 3.335 |  |  | Increase | Clearance |
|  |  | EEG Beta Power (REM) | -0.041 | 0.806 | 0.734 | 1.090 |  |  | Increase | Production |
|  | Negative | R<sub>p</sub> | 0.963 | -0.255 | -0.341 | -0.320 | -0.295 | -0.161 | Increase | Clearance |
|  |  | PTT (NREM) | -0.058 | 0.019 | 0.025 | 0.024 | 0.022 | 0.012 | Increase | Clearance |
|  |  | HRV (NREM) | 0.114 | -0.043 | -0.058 | -0.054 | -0.050 | -0.027 | Increase | Production |
|  |  | EEG Delta Power (NREM) | -0.041 | 0.014 | 0.019 | 0.018 | 0.017 | 0.009 | Increase | Clearance |
|  |  | EEG Theta Power (NREM) | 0.169 | -0.044 | -0.059 | -0.056 | -0.051 | -0.028 | Increase | Production |
|  |  | EEG Theta Power (REM) | 0.087 | -0.027 | -0.036 | -0.034 | -0.031 | -0.017 | Increase | Production |
|  |  | EEG Beta Power (REM) | -0.135 | 0.072 | 0.096 | 0.090 | 0.083 | 0.045 | Increase | Production |
| Sleep Condition *Hypno<sub>s</sub>* | Positive | NREM Duration | -0.182 | 0.263 | 0.257 | -0.143 |  |  | Increase | Clearance |
|  |  | REM Duration | -0.519 | 0.204 | 0.200 | -0.111 |  |  | Increase | Production |
|  |  | N1 Duration | 0.834 | -0.267 | -0.262 | 0.145 |  |  | Increase | Clearance |
|  |  | WASO Duration | 0.044 | -0.022 | -0.021 | 0.012 |  |  | Increase | Clearance |
|  | Negative | NREM Duration | -0.090 | 0.032 | 0.059 | 0.049 | 0.042 | -0.003 | Increase | Clearance |
|  |  | REM Duration | 0.525 | -0.069 | -0.128 | -0.106 | -0.092 | 0.006 | Increase | Clearance |
|  |  | N1 Duration | 0.801 | -0.076 | -0.141 | -0.116 | -0.101 | 0.007 | Increase | Clearance |
|  |  | WASO Duration | 0.273 | -0.019 | -0.036 | -0.030 | -0.026 | 0.002 | Increase | Clearance |
| Sleep Deprivation/ Wake *Physio<sub>w</sub>* | Positive | R<sub>p</sub> | 0.621 | 0.149 | 0.180 | -1.624 |  |  | Increase | Clearance |
|  |  | PTT | 0.190 | 0.054 | 0.065 | -0.583 |  |  | Increase | Production |
|  |  | HRV | -0.242 | -0.065 | -0.079 | 0.712 |  |  | Increase | Clearance |
|  |  | EEG Delta Power | -0.487 | -0.112 | -0.135 | 1.223 |  |  | Increase | Production |
|  |  | EEG Theta Power | -0.382 | -0.102 | -0.124 | 1.116 |  |  | Increase | Production |
|  |  | EEG Beta Power | 0.371 | 0.114 | 0.138 | -1.246 |  |  | Increase | Clearance |
|  | Negative | R<sub>p</sub> | 0.626 | -0.300 | -0.301 | -0.129 | -0.129 | -0.106 | Increase | Clearance |
|  |  | PTT | -0.004 | 0.002 | 0.002 | 0.001 | 0.001 | 0.001 | Increase | Clearance |
|  |  | HRV | 0.136 | -0.094 | -0.094 | -0.040 | -0.041 | -0.033 | Increase | Production |
|  |  | EEG Delta Power | -0.658 | 0.335 | 0.337 | 0.144 | 0.145 | 0.118 | Increase | Production |
|  |  | EEG Theta Power | -0.391 | 0.211 | 0.212 | 0.091 | 0.091 | 0.074 | Increase | Production |
|  |  | EEG Beta Power | 0.062 | -0.032 | -0.032 | -0.014 | -0.014 | -0.011 | Increase | Clearance |

\**R<sub>p</sub>*, Parenchymal Resistance; PTT, Pulse Transit Time; HRV, Heart Rate Variability; EEG, Electroencephalography; *Physio<sub>s</sub>* and *Hypno<sub>s</sub>*, single predictors for sleep and *Physio<sub>w</sub>* single predictors for sleep deprivation.

**Supplemental Table 9.**
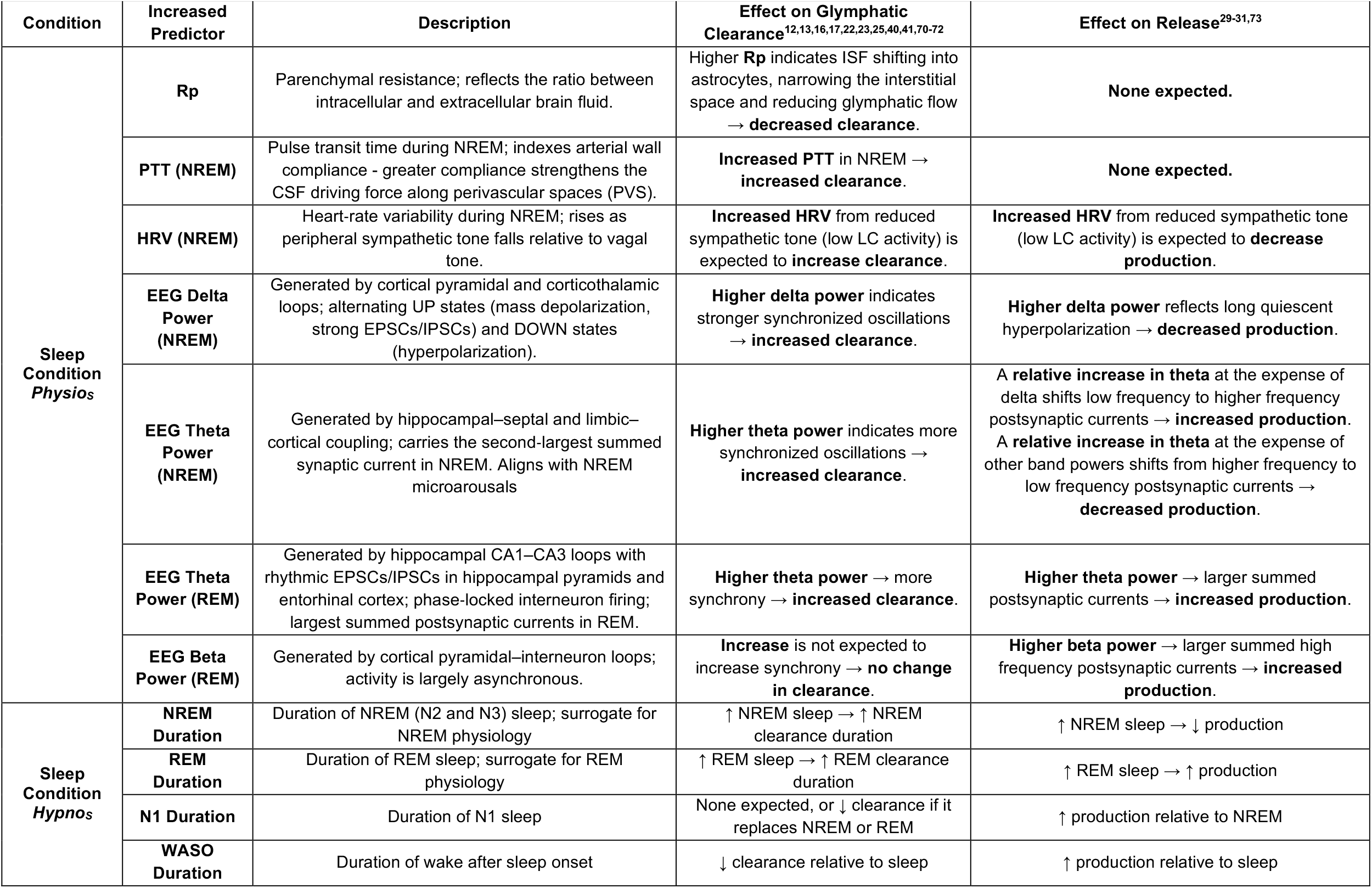
Effects of increasing predictors in *Physio*_*S*_ and *Hypno*_*S*_ on sleep-dependent glymphatic clearance and synaptic-metabolic solute production.

**Supplemental Table 10.**
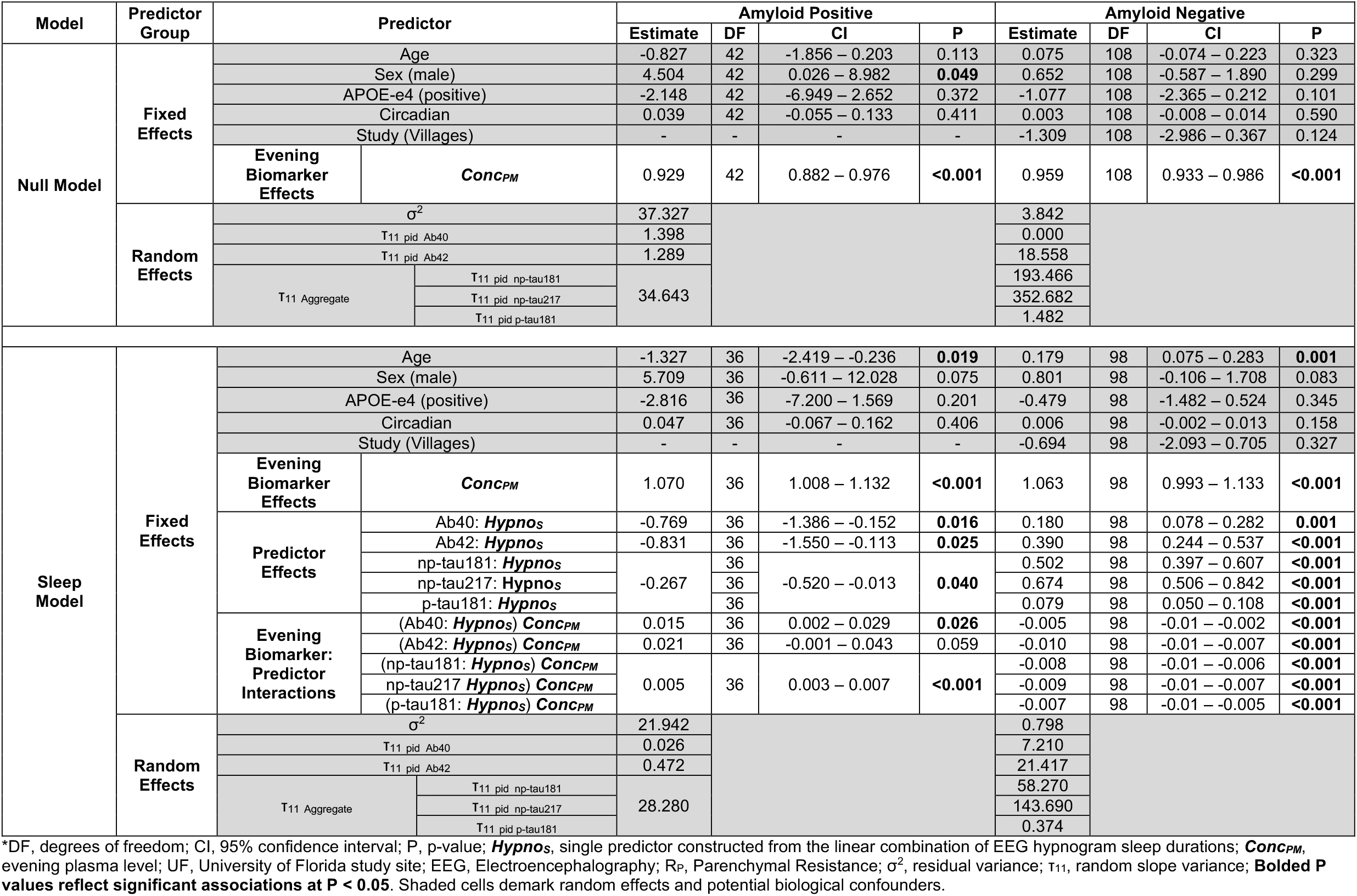
Prediction of morning plasma AD biomarker levels following overnight sleep using the *null model* and the *neuro-glymphatic model* with *Hypno*_*S*_*.

**Supplemental Table 11.**
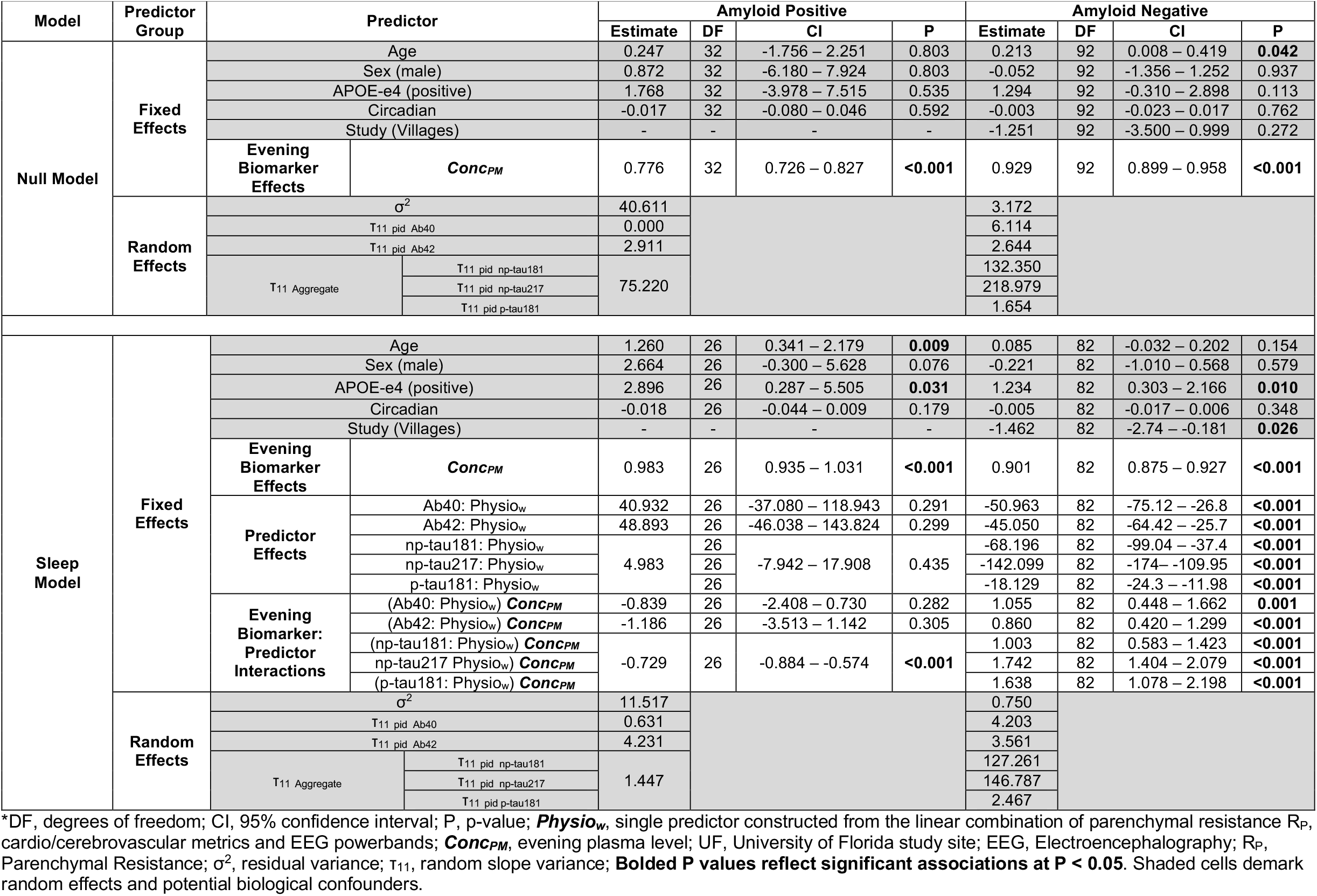
Prediction of morning plasma Aβ and tau levels following overnight sleep using the *null model* and the *neuro-glymphatic model* with *Physio*_*w*_*.

**Supplemental Table 12.**
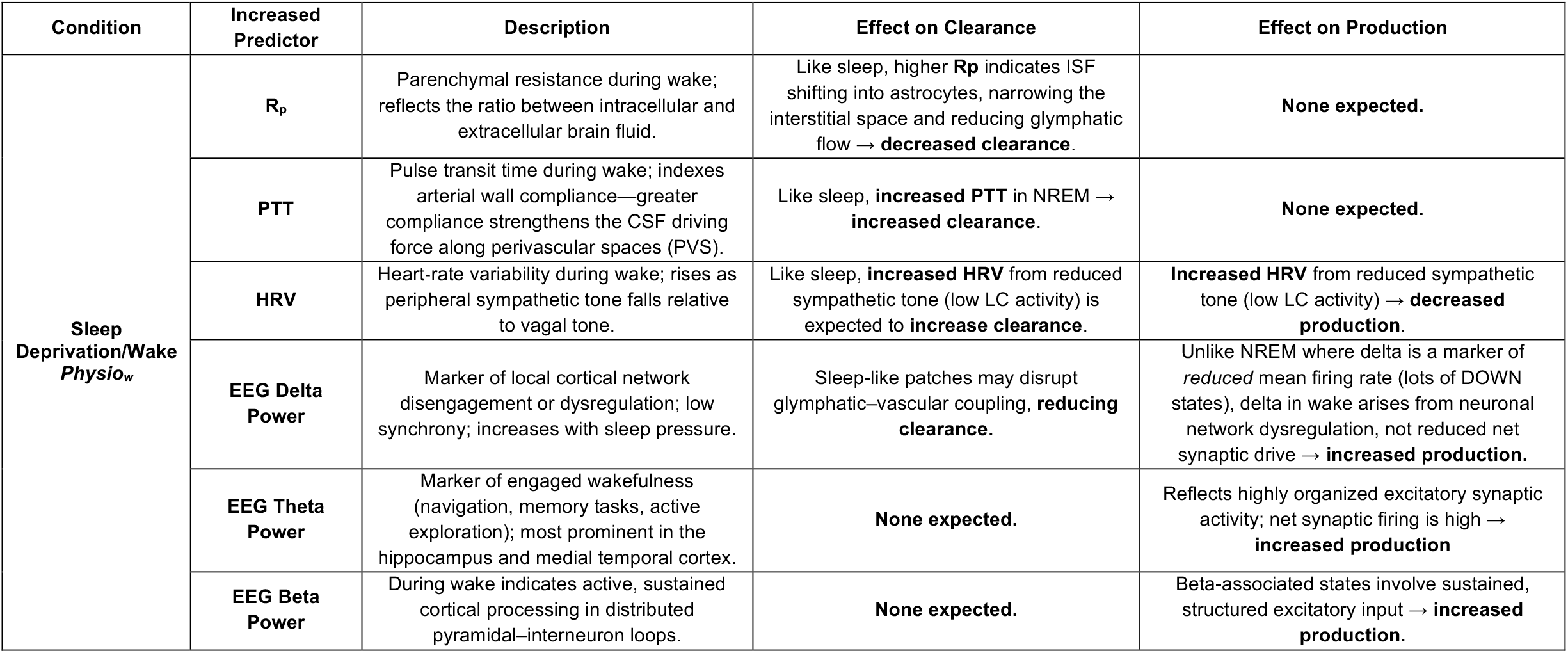
Effects of increasing predictors in *Physio*_*W*_ on glymphatic clearance and synaptic-metabolic solute production during sleep deprivation.

## Data availability

The data supporting the results in this study are available from the corresponding author with Institutional Review Board approval and Data Use Agreement permitting non-commercial use of data for independent validation, publication and sharing of new findings. For participants with one or more measures that failed the pipeline processing quality control, we will, upon special request, provide the measures that passed quality control, along with the unprocessed sensor data for the others where possible.

## Code availability

Code used for the analysis and to produce the figures will be available on Zenodo. The code for the compartment model simulations is available on github.com^75^.

